## Supplementary Material for "Phenotypic and Genetic Characteristics of Retinal Vascular Parameters and their Association with Diseases"

### Data

Supplementary File 1 - GWAS top hits (UKBB) before pruning [1]

Supplementary File 2 - IDP Gene scores (UKBB) [2]

Supplementary File 3 - PascalX IDP-IDP cross gene scores [3]

Supplementary File 4 - PascalX IDP-disease cross gene scores [4]

Supplementary File 5 - GWAS top hits (RS) before pruning [5]

Supplementary File 6 - IDP Gene scores (RS) [6]

Supplementary File 7 - SNPs replication: UKBB replicated in RS, and RS replicated in UKBB [7]

Supplementary File 8 - Genes replication: UKBB replicated in RS, and RS replicated in UKBB [8]

Supplementary Table 1 - Significant genes per IDP [9]

Supplementary Table 2 - Significant pathways per IDP

Supplementary Table 3 - Phenotypic and genetic correlations, discovery and replications

Supplementary Table 4 - Diseases information

Supplementary Table 5 - Significant genes per diseases

Supplementary Table 6 - Significant pathways per diseases

Supplementary Table 7 - DRIVE temporal angles

Supplementary Table 8 - MR risk factors (IVW)

Supplementary Table 9 - MR risk factors (all methods)

Supplementary Table 10 - MR risk factors (Egger intercept)

Supplementary Table 11 - MR risk factors (LOO)

Supplementary Table 12 - IDPs phenotypic association with diseases

---

[1] https://drive.google.com/file/d/1GMLRNraNw_SJO9XxeX6LNaD6n86SsOIz/view?usp=sharing
[2] https://drive.google.com/file/d/1xlVq1bSMJy6N9HDh9eToRmshzVKfCbSx/view?usp=sharing
[3] https://drive.google.com/file/d/1Jtujale4iz_MIIj38sbc-423xGV5M2Wr/view?usp=sharing
[4] https://drive.google.com/file/d/1jHmReJ5QJ_EY-xW2xZ9uV-JPdWFVV8h_/view?usp=sharing
[5] https://drive.google.com/file/d/11428BJTl60ZEcqveTT2oxacwjUbQjsj7/view?usp=drive_link
[6] https://drive.google.com/file/d/1X8HneGYsfsQx4N89VVQ-4wCSNZPVL_xt/view?usp=drive_link
[7] https://docs.google.com/spreadsheets/d/192Iew0WUHMaIG2w4jDJ5DtfY8M014_uCzpGnWl0YZhc/edit?usp=sharing
[8] https://drive.google.com/file/d/1soq_Pw6zu6DFel04yoeMQDWHcGIJxixL/view?usp=drive_link
[9] https://docs.google.com/spreadsheets/d/1vS11c0Z0aUeXtDBw396npgwNBpIzLa6gFxIl2NC7sWw/edit#gid=0

### 1 Image segmentation

#### 1.1 Vessel segmentation

Descriptions of ARIA and LWNET image segmentation methods, as well as their vessel segment-wise artery-vein classification accuracy are given in Supplemental Text 4 of our previous work on retinal tortuosity [1]. We made a small modification to that method for this study: ARIA is now used solely for skeletonization and the identification of diameters. It newly skeletonizes pixel-wise masks coming from LWNET instead of its own masks.

#### 1.2 Optic disc segmentation

We manually annotated 100 Optic Discs (ODs) from Color Fundus Images (CFIs) of 100 UK Biobank (UKBB) participants, creating an in-house ground truth dataset. We then retrained a previously published CNN [2], using 80 random ground truth images for training, and the remaining 20 images for validation. Using this model, we achieved an average DICE score of 0.86 on validation (Fig. 1).

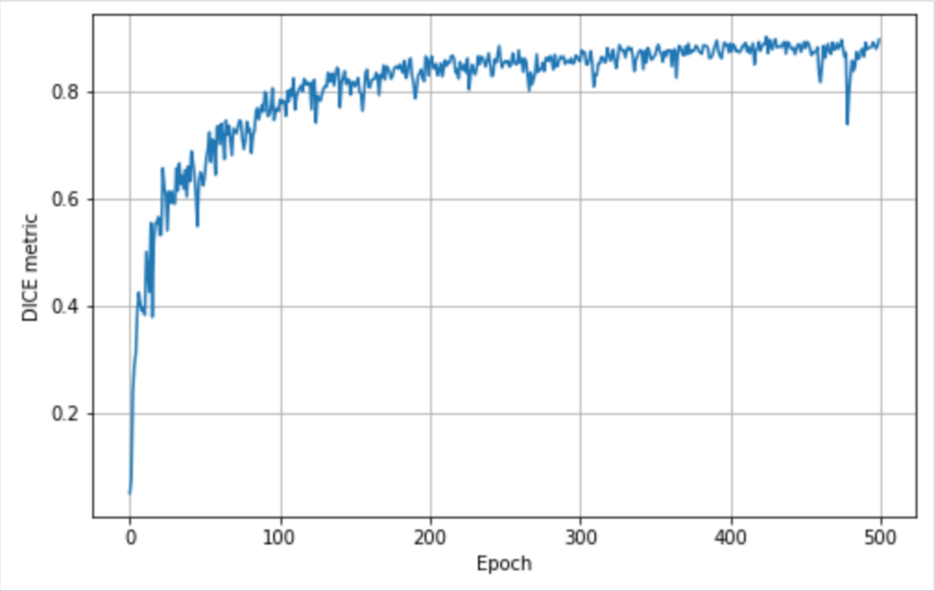


Figure 1: Performance of re-trained CNN on the validation set.

### 2 Methods for phenotype extraction

We implemented various methods to measure retinal vascular phenotypes, including diameter variability, central retinal equivalent, the major temporal angle, the number of bifurcations, fractal dimension, vascular density, and tortuosity. These measurements were extracted for arteries, veins, and without vessel type differentiation whenever feasible. In this section, we describe the methods used.

Fig. 2 showcases visualizations of some of these methods applied to a CFI of the public DRIVE-fundus-dataset, following the methodology used in the UKBB.

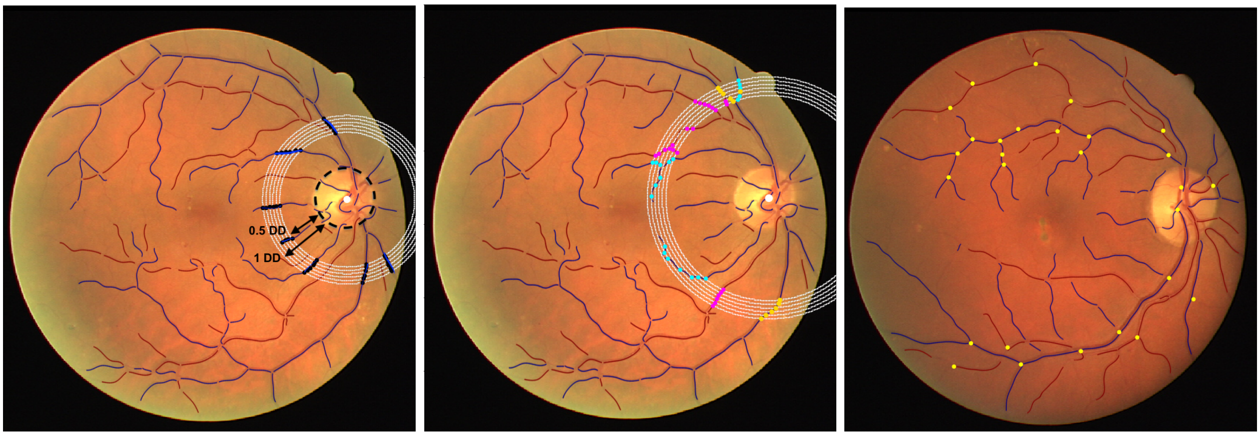


(a) V central retinal equivalent (b) V temporal angle (c) Number of bifurcations

Figure 2: Visualization of some IDPs on a DRIVE CFI: **a)** venular central retinal equivalent, **b)** venular temporal main angle, and **c)** number of bifurcations.

#### 2.1 Median diameter and diameter variability

To compute the median diameter, we utilized ARIA tool [3] to obtain the diameter value per centerline pixel, as well as the assignment of these pixels to vessel segments. We then calculated the median diameter separately for arteries ('A Diameter') and veins ('V Diameter'). This was done by first dividing the data by vessel segment, computing the median diameter values for each segment, and then taking the median across all the segments. Additionally, we computed the overall median diameter without differentiation by vessel type ('Diameter'), considering all the diameter values. See equation 1.

$$\begin{aligned} \text{`A Diameter'} &\equiv \text{median arteries diameter} = median(median(\vec{d_{A\,segment}})) \\ \text{`V Diameter'} &\equiv \text{median veins diameter} = median(median(\vec{d_{V\,segment}})) \\ \text{`Diameter'} &\equiv \text{median diameter} = median(median(\vec{d_{segment}})) \end{aligned} \tag{1}$$

Where $\vec{d_{A\,segment}}$ refers exclusively to the arterial diameters within a segment, $\vec{d_{V\,segment}}$ refers to the veins diameters within a segment, and $\vec{d_{segment}}$ represents all diameter values within a segment, including both arteries and veins.

To assess the diameter variability, we employed both a dimensional and an adimensional approach. The dimensional one uses the standard deviation formula, std, equation 2, while the adimensional uses the coefficient of variation regarding the median, CVMe, equation 3.

$$\begin{aligned}
&\text{`A std Diameter'} \equiv \text{standard deviation arteries diameter} = std(\vec{d_A}) \\
&\text{`V std Diameter'} \equiv \text{standard deviation veins diameter} = std(\vec{d_V}) \\
&\text{`std Diameter'} \equiv \text{standard deviation diameter} = std(\vec{d})
\end{aligned} \tag{2}$$

Here, $\vec{d_A}$ represents the values of all the arterial diameters, $\vec{d_V}$ the vein diameters, and $\vec{d}$ represents all the diameter values, both arteries and veins. Note that no segment filter is applied here.

$$\begin{aligned}
&\text{`A CVMe Diameter'} = \frac{D_{median}(\vec{d_A})}{\mid median(\vec{d_A}) \mid} \\
&\text{`V CVMe Diameter'} = \frac{D_{median}(\vec{d_V})}{\mid median(\vec{d_V}) \mid} \\
&\text{`CVMe Diameter'} = \frac{D_{median}(\vec{d})}{\mid median(\vec{d}) \mid}
\end{aligned} \tag{3}$$

Where $D_{median}(x)$ is the deviation from the median.

#### 2.2 Central retinal equivalents

There are two central retinal equivalents: the central retinal arterial equivalent (‘A central retinal eq’, also known as ‘CRAE’), and the central retinal venular equivalent (‘V central retinal eq’, also known as ‘CRVE’). The methodology for measuring them in this study is based on [4]. This method assumes that the OD is placed in the center of the image, and so, both the temporal and nasal vessels are visible. According to this method, to get the central retinal arterial equivalent, it measures the OD position, the OD diameter, and at a distance $r$ of 0.5 OD diameter, it selects the 6 arteries (if measuring ‘A central retinal eq’) at that distance, i.e. we get a list of diameters like $[d_r^1, d_r^2, d_r^3, d_r^4, d_r^5, d_r^6]$. Of these six arteries, the ones of interest are the one with the largest diameter, $d_r^1$, and the one with the smallest diameter, $d_r^6$. Then, the procedure tracks these two arteries from a distance $r$ of 0.5 OD to 1 OD of the OD center and computes their median diameter. So we end up with two values: $d_{Amax} = median(d_r^1)$, $d_{Amin} = median(d_r^6)$, and from here we compute the central retinal arterial equivalent applying equation 4a. Similarly, the central retinal venular equivalent selects veins instead of arteries and uses equation 4b.

However, the UKBB CFIs have the OD on one side and only the temporal vessels are visible, so instead of six we select the three main arteries or veins. To this end, we analyzed two alternative methods. The first one is identical to the one outlined above, with the only difference that we select the three main vessels instead of six (‘central retinal eq2’), i.e. we select $d_r^1$, and $d_r^3$ instead of $d_r^1$, and $d_r^6$. The potential problem with this method is that if there is a vessel discontinuity for the main vessel, the method does not work. Therefore, we propose an alternative method to deal with such situations ('central retinal eq'). In this method, we similarly get the vessel with the highest diameter and the one with the third highest at 0.5 OD. But, we apply the equation 4 over these two diameters, so $d_{Amax} = d_r^1$ and $d_{Amin} = d_r^3$ are not the median, but the diameter values at these particular distances. We obtain these two parameters at distances $r$ between 0.5 to 1 OD and we apply again the previous formula for each radius. We then compute the median of the central retinal equivalent that we get for each $r$ (see algorithm 1). The advantage of this second method is that the measure is well-defined even if there is a discontinuity in the main vessels, so can be obtained for more images, and for this reason, we use it as the one of the main IDPs. However, we observed that the values obtained from both methods are highly correlated, phenotypically and genetically, $r^{(p)}$=0.77 and $r^{(g)}$=0.98 for the arteries, and $r^{(p)}$=0.80 and $r^{(g)}$=1.00 for the veins (Fig. 3).

$$\begin{aligned} \text{`A central retinal eq'} &= 0.88 * ({d_{Amin}}^2 + {d_{Amax}}^2)^{1/2} \\ \text{`V central retinal eq'} &= 0.95 * ({d_{Vmin}}^2 + {d_{Vmax}}^2)^{1/2} \end{aligned} \tag{4}$$

**Algorithm 1** Central retinal equivalent algorithm

```
for image in dataset do
  Measure OD position
  if OD position found then
    list_radius = [0.5 OD, 0.6 OD, 0.7 OD, 0.8 OD, 0.9 OD, 1.0 OD]
    for r in list_radius do
      Plot circle with center at OD and radius = r
      Search the intersections between the circle and vessels
      if type=='A central retinal eq': then
        Filter by arteries
        cte = 0.88
      if type=='V central retinal eq': then
        Filter by veins
        cte = 0.95
      Select the 3 vessels with the biggest diameter
      Of these 3 values select the biggest and the smallest ones, i.e. d_max and d_min
      CRE(r) = cte*(d_min^2 + d_max^2)^(1/2)
      return median(CRE(r))
```

#### 2.3 Main temporal angles

The methodology here presented is an automated version based on the semi-automatic process explained in [5], and adapted to images with the OD located on one side. For that reason, we do not measure the nasal angles, but only the temporal angles. We measured the OD position and plotted a circle at a radius, $r_0$, of 240 pixels, (this value is around 1/5 of the diameter of the CFIs and needs to be adapted to the image size, e.g. for the DRIVE images $r_0$=120 pixels). Then we select the intersections between the circle and the vessels. We filter by the vessel type of interest, arteries if 'A temporal angle' and veins if 'V temporal angle'. And to select the ones in the major branches we select the ones with the biggest diameters values and that fulfill certain requirements (the angles between the two points have to make an angle, with origin in the OD center, bigger than 75° and smaller than 200°. These values were decided based on the input from an expert opthalmologist, suggesting that values outside this range are likely to be artifacts. We select the angle between these two points as the value of the temporal angle for radius $r = r_0$. However, to have a more robust technique, this process is repeated at six different radii (i.e. $r \in [r_0, r_0 + \delta, r_0 + 2\delta, r_0 + 3\delta, r_0 + 4\delta, r_0 + 5\delta]$, where we used $\delta$=10 for the UKBB and 7 for DRIVE) values and applied a "majority vote" (with an interval of error) to decide if the measure is valid or not, algorithm 2.

**Algorithm 2** Temporal angles algorithm

```
for image in dataset do
  Measure OD position
  if OD position found then
    r_0 = 240 {Initial radius value}
    δ = 10 {Increment value}
    list_radius = [r_0, r_0 + δ, r_0 + 2δ, r_0 + 3δ, r_0 + 4δ, r_0 + 5δ]
    for r in list_radius do
      Plot circle with center at OD and radius = r
      Search the intersections between the circle and vessels
      if type=='A temporal angle' then
        Filter by arteries

      if type=='V temporal angle' then
        Filter by veins

      Compute the angles between each two points
      Select the points with the biggest diameter and whose angle is > 75 and < 200
      angle_value(r)

    Compute majority vote between the different angle_value(r)
    if Majority votes agree then
      return mean(angle_value(r))

  else
    Not possible to compute the angle for this image
```

To test for potential dependence on the values of $r_0$, $\delta$, and the angles range error values, different choices close to these numbers were tested, showing similar results independently of small differences in these numbers.

#### 2.4 Number of bifurcations

This phenotype has been extensively studied in the literature [6, 7, 8]. Its accurate measurement is a complex problem, and both DL and classical image analysis are used. However, to the best of our knowledge, for none of these methods their code was openly accessible at the moment of starting our analysis. For this reason, we decided to develop a simple method based on our vessel segmentation tools. The LWNET algorithm [9] provides artery-vein segmentation, while ARIA [3] allows us to enumerate each segment in the vasculature. To measure the number of bifurcations, we initially attempted to identify cases where two segment endpoints of the same type (either arteries or veins) were proximal to the endpoint of a third segment. To achieve this, we needed to define a radius defining a proximity neighborhood. However, we found that this methodology excluded many actual bifurcations because two endpoints often appeared close to each other while the third endpoint was farther away. Since increasing the proximity radius would potentially lead to false positives, we devised a modified approach. We examined all endpoints to determine if there was just one additional endpoint of the same vessel type in the neighborhood. Subsequently, we removed bifurcations that were too close to each other as they were considered instances of overcounting. This adjustment aimed to strike a balance between detecting true bifurcations and minimizing false positives, algorithm 3.

**Algorithm 3** Number of bifurcations algorithm

```
for image in dataset do
  Select the end-points for each vessel segment and save the vessel type of the corresponding segment
  for each end-points do
    Search end-points in the neighborhood of a distance = Cte_N1
    if they are the same type of vessel then
      Add this point as a bifurcation
    Remove bifurcation points in a neighborhood of a distance = Cte_N2
    return Number of bifurcations
```

Due to the potential dependence of the output on the values of the two neighborhood threshold values ($Cte_{N1}$, $Cte_{N2}$), different values close to these numbers were tested, showing similar results independently of small differences in these numbers.

#### 2.5 Vascular density

We computed vascular density as the fraction of pixels annotated as vessels within the fundus mask area.

**Algorithm 4** Vascular density

**Input:** Pixel-wise artery-vein masks
To compute overall VD, provide all 3 color channels, to compute arterial VD provide red channel, or to compute venular VD, provide a blue channel

Remove all pixels outside the fundus mask from analysis

Return

$$VD = \frac{\#\ non - black\ pixels}{\#\ total\ pixels}$$

#### 2.6 Fractal dimension

We implemented the Minkowski–Bouligand dimension—also known as box-counting dimension—to quantify the fractal dimension (FD) of retinal vessels. Analytically, for any surface $S$, it is defined as:

$$dim_{box}(S) = \lim_{\epsilon \to 0} \frac{\log(N(\epsilon)}{\log(1/\epsilon)}$$

To estimate this quantity, we counted the number of boxes ($N$) required to fully cover the vasculature across a set of scales ($\epsilon$), such that the boxes at the largest scale are wider than the widest blood vessels in UKBB images ($\approx$ 32 pixels). FD is then estimated as minus the slope of the regression $\log N \sim \log \epsilon$.

**Algorithm 5** Fractal dimension

**Input:** Gray-scale, pixel-wise artery-vein masks
To compute arterial FD, provide only the red channel, to compute venular FD, provide only the blue channel

Define a set of scales bounded between $2^{0.01}$ and $2^5$, and equally spaced in log scale

At each scale, count the number of boxes required to cover vasculature

Regress $\log \epsilon$ onto $\log N$

Return slope

#### 2.7 Tortuosity

We defined all implemented tortuosity measurements in Supplemental Text 2 of our previous work on tortuosity [1]. The numeric labels remain the same.

#### 2.8 Ratios

The ratio of a phenotype was computed as the phenotypes of the arteries over the phenotype of the veins, equation 5.

$$
\begin{aligned}
\text{`ratio tortuosity'} &= \frac{\text{`A tortuosity'}}{\text{`V tortuosity'}} \\
\text{`vascular density'} &= \frac{\text{`A vascular density'}}{\text{`V vascular density'}} \\
\text{`ratio median diameter'} &= \frac{\text{`A median diameter'}}{\text{`V median diameter'}} \\
\text{`ratio central retinal eq'} &= \frac{\text{`A central retinal eq'}}{\text{`V central retinal eq'}}
\end{aligned}
\tag{5}
$$

### 3 Previous associations with diseases

The main temporal angles of arteries and veins (‘A temporal angle’, ‘V temporal angle’) have been linked to diseases such as severe myopia, retinopathy of prematurity (ROP), and proliferative diabetic retinopathy (PDR) [10, 5].

The vessel diameter for arteries, veins, and their ratio (‘A median diameter’, ‘V median diameter’, ‘ratio median diameter’) have been linked to diseases such as hypertension and coronary heart disease risk [11, 12, 13, 14, 15, 16, 17, 18, 19, 20].

The central retinal equivalent of the arteries, veins, and their ratio (‘A central retinal eq’, ‘V central retinal eq’, ‘ratio central retinal eq’) have been linked with diseases such as hypertension [4].

The diameter variability of the arteries and veins (‘A std diameter’, ‘V std diameter’) have been linked with diseases such as hypertension and diabetes [18].

The number of bifurcations (‘bifurcations’) have been linked to diseases such as hypertension [21].

The tortuosity of veins, arteries and their ratio (‘A tortuosity’, ‘V tortuosity’, ‘ratio tortuosity’) have been linked to diseases such as cardiovascular risk factors [22, 23].

The vascular density of the arteries, veins, and their ratio (‘A vascular density’, ‘V vascular density’, ‘ratio vascular density’) have been linked to diseases such as stroke, coronary artery disease, and AD [24, 25].

### 4 Correlation structure and heritabilities of extended list of retinal vascular IDPs

Although the main paper focuses on the examination of 17 IDPs, numerous additional IDPs were computed, often representing alternative measurements of similar traits. This section shows the phenotypic and genetic correlation between the complete list of IDPs, their SNP heritabilities (Fig. 3, and 4), as well as the linear and logistic associations between all IDPs and various risk factors and diseases (Suppl. Text 19), after correcting for the covariates (Fig. 5).

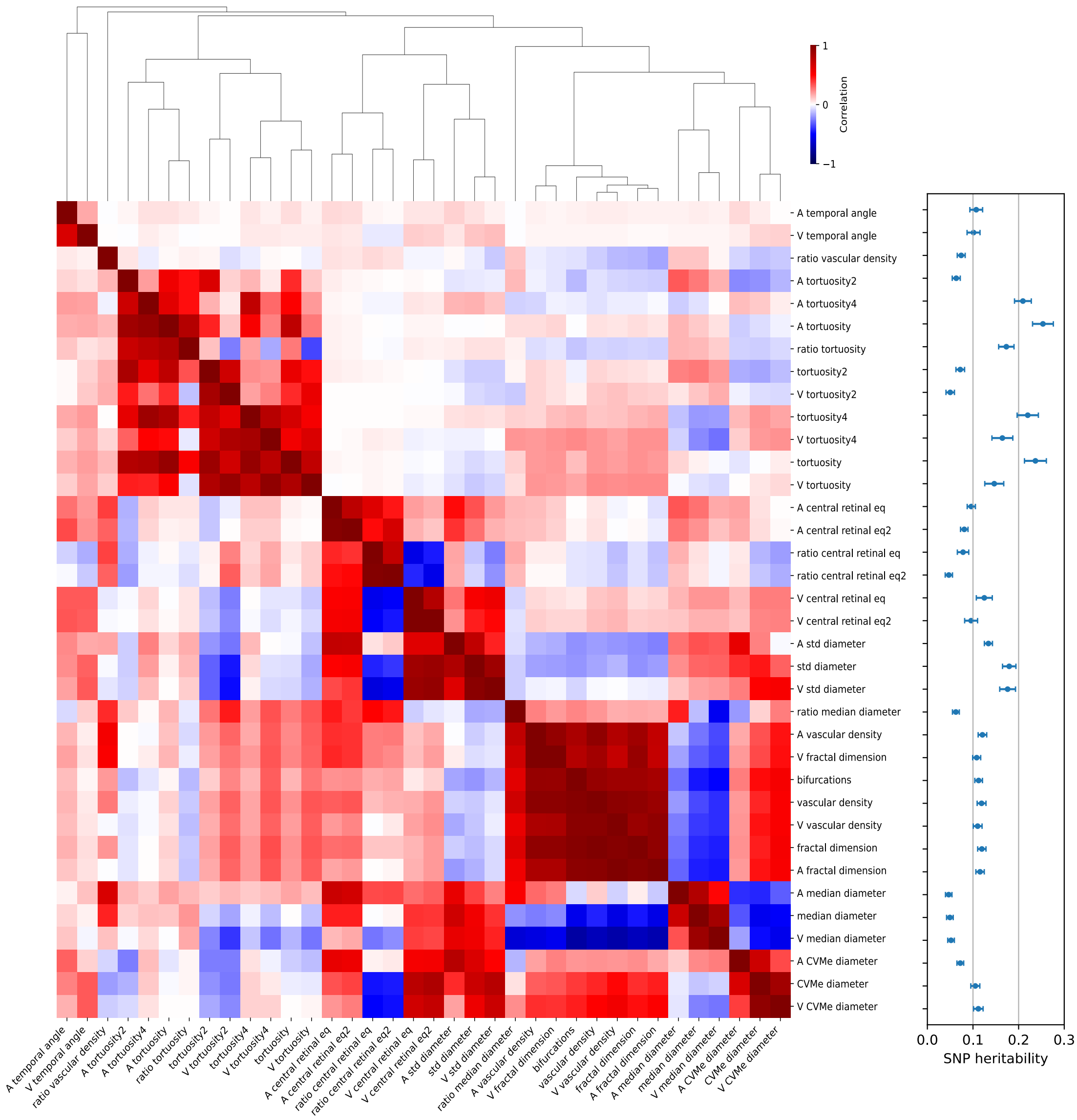


Figure 3: **Left:** Phenotypic, genetic correlations and heritability of an extended list of 36 retinal vascular IDPs. The upper half of the square represents the phenotypic correlation, whereas the lower half represents the genetic correlation. **Right:** SNP heritabilities of the 35 IDPs. Both plots were computed after subtracting the effects of the covariates. The covariates utilized were those employed for the main IDPs.

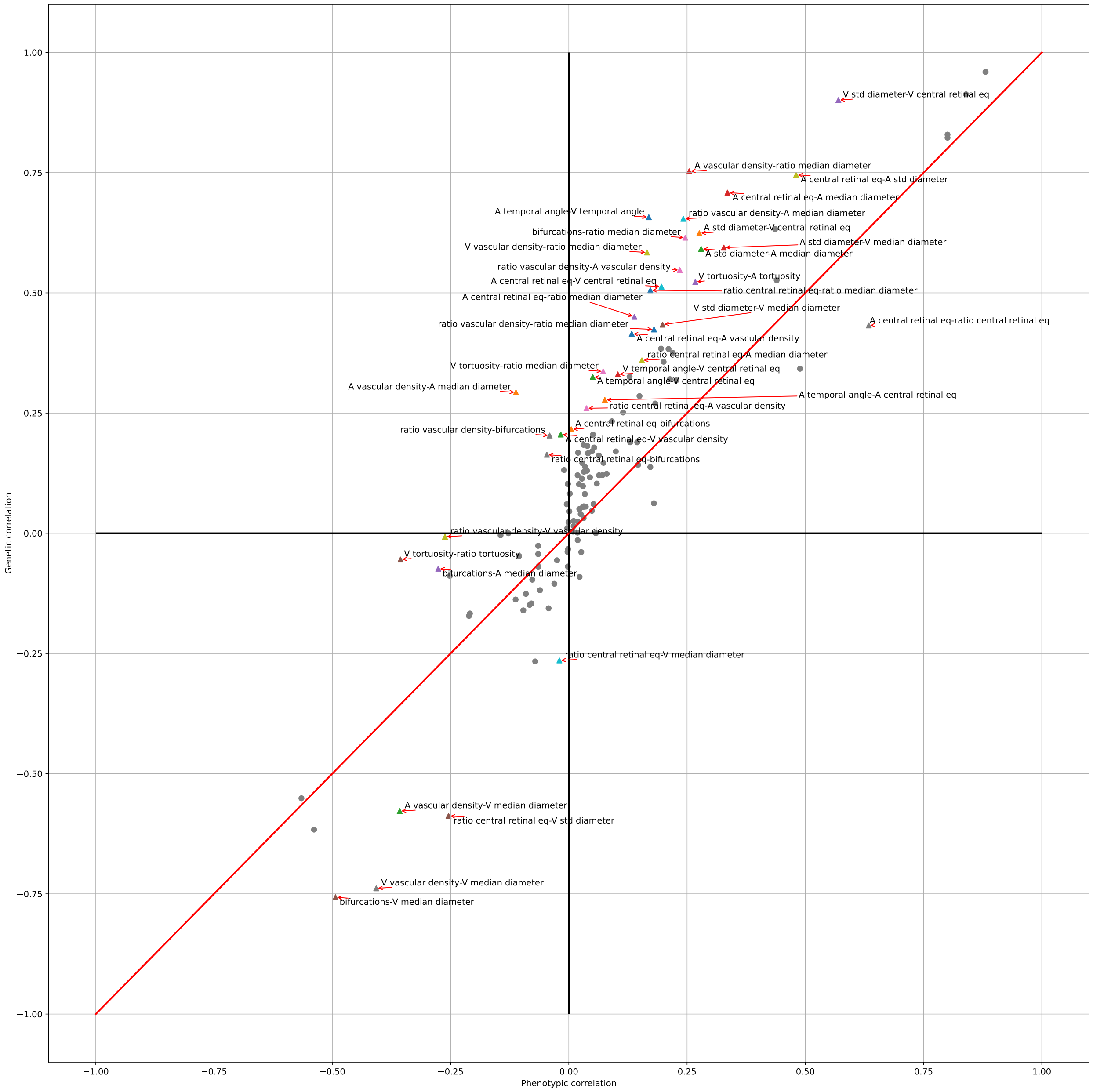


Figure 4: Scatter plot illustrating the phenotypic-genetic correlation across the entire list of IDPs. Phenotype pairs with $|r^g - r^p| > 0.2$ are labeled. The covariates utilized were those employed for the main IDPs.

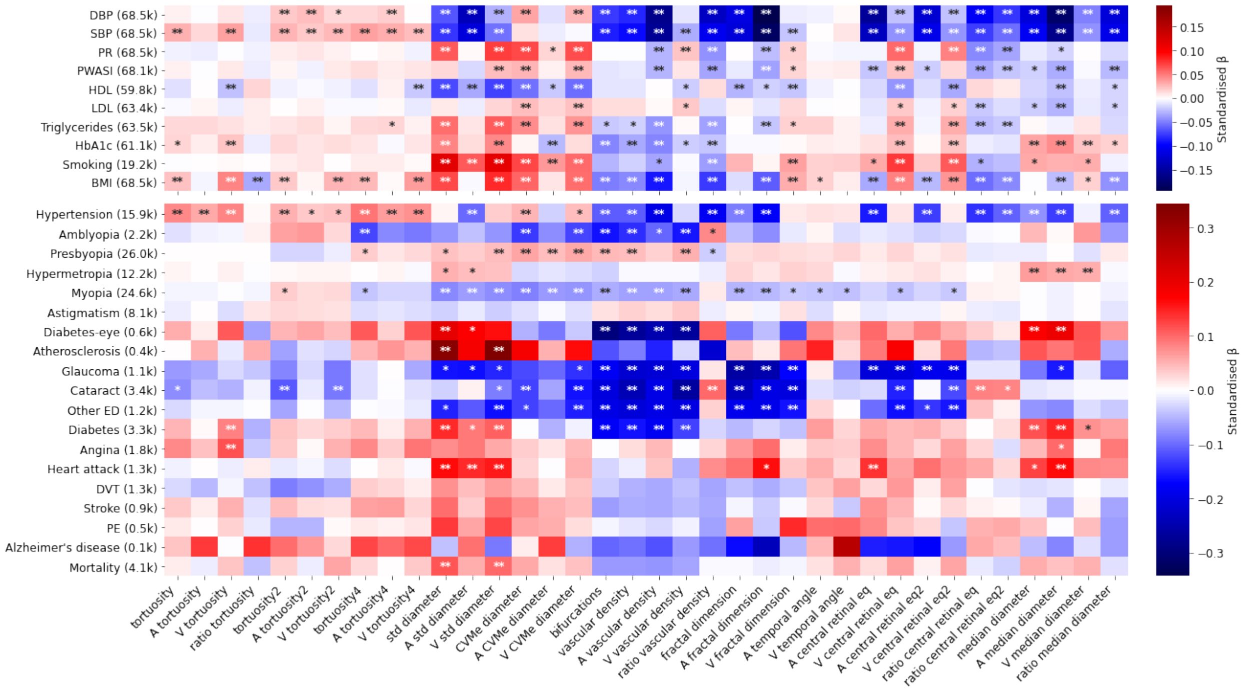


Figure 5: Phenotypic association of IDPs with risk factors and diseases. The x-axis shows IDPs and the y-axis shows risk factors and diseases. For risk factors, the numbers in parentheses correspond to the maximum sample size of subjects with disease information, and for diseases to the number of cases. Linear **(upper)** and logistic **(lower)** regressions were used for continuous and binary diseases respectively. In both models, IDPs were corrected for age, sex, eye geometry, batch effects, and ethnicity. The color indicates standardized effect sizes, linear and logistic regressions. The asterisks indicate level of statistical significance ($* : p < 0.05/N_{test}$, $** : p < 0.001/N_{test}$, being $N_{test} = N_{IDPs} \times N_{Diseases}$). Labels: 'PR': Pulse Rate, 'PWASI': Pulse wave Arterial stiffness index, 'HDL': High-Density Lipoprotein, 'HbA1c': Glycated hemoglobin, 'Alcohol': Alcohol intake frequency, 'Smoking': pack-years, 'Diabetes-eye': diabetes related to the eye, 'DVT': Deep Vein Thrombosis, 'Other ED': all types of severe eye diseases not included explicitly, 'PE': pulmonary embolism.

### 5 Distribution of retinal vascular IDPs

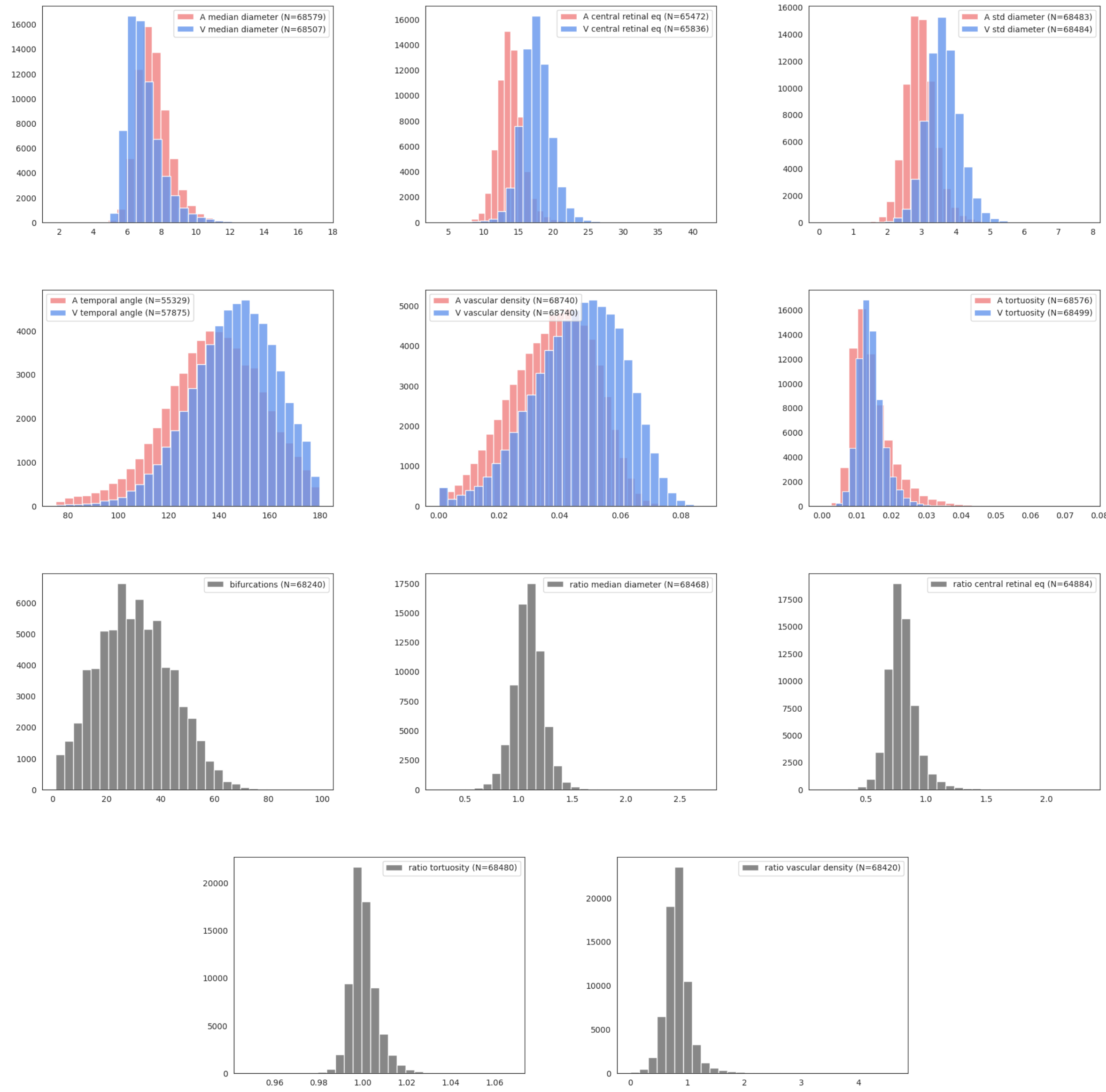


Figure 6: UKBB raw distributions of the 17 main IDPs per subject. Outlier values, defined as $|\text{value}| > mean + 10std$), were removed. This outlier removed was especially important for the 'ratio vascular density'.

#### 5.1 Principal Component Analysis

In addition to analyzing the 17 main IDPs separately, we applied component reduction techniques to see if any of these PCs encapsulated the information of the individual IDPs better. In this section, you can find the comparison of these PCs with the IDPs (Fig. 7), as well as the association of the PCs with the diseases and risk factors we are studying (Fig. 8).

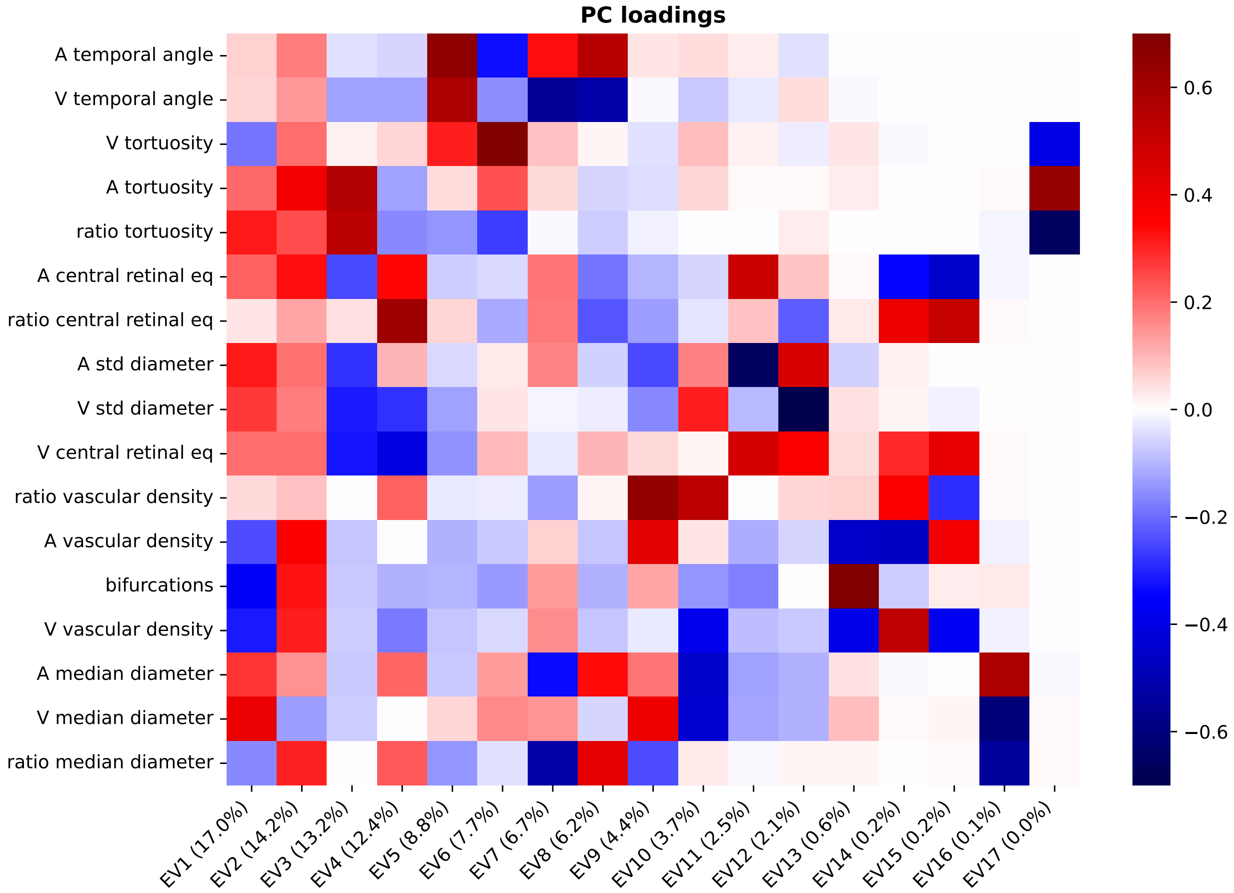


Figure 7: Eigenvector mappings on retinal IDPs. Brackets show the percentage of variance explained by each PC. The cumulative variance of the first 9 PCs surpasses 90%. PCs 1 and 2 predominantly reflect variations in vascular sparsity and vascular density. PC3 combines elevated arterial tortuosity with low diameter variability. PC4 reflects a unbalance between the diameters of the central retinal artery and vein, while PC5 is characterized by their temporal angles.

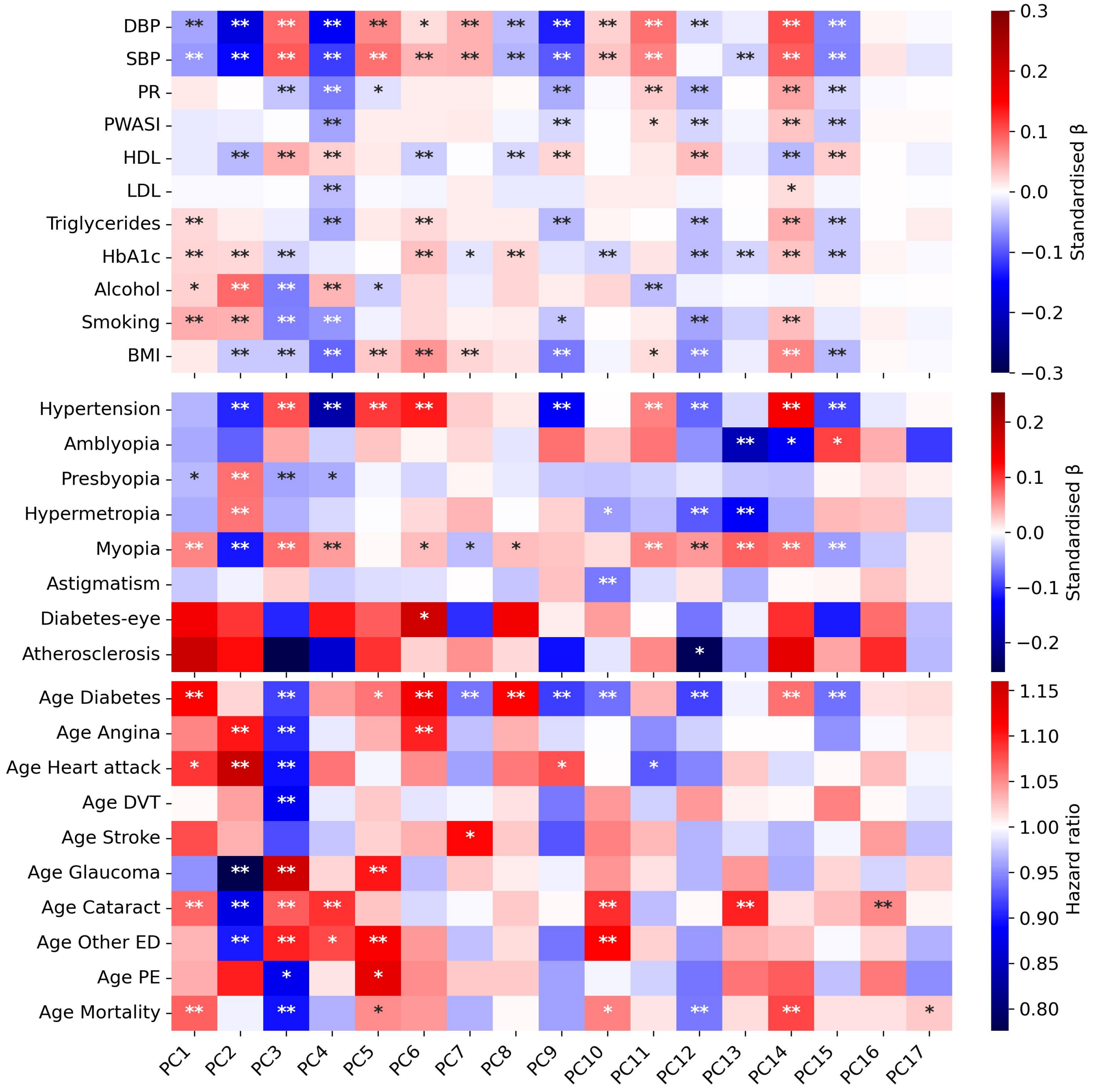


Figure 8: Principal Component Analysis (PCA) of retinal IDPs revealed disease associations on a scale comparable to that of the initial IDPs, as depicted in Figure 4 in the main manuscript.

### 6 Validation of IDPs

To validate the phenotypes, two procedures were used. In the UKBB, multiple random images were visually inspected in different quantiles to search for potential confounders in the measurements. Some phenotypes suspected to be affected by this bias were refined if possible or removed. Additionally, another validation was performed for the number of bifurcations and the temporal angles.

Although they are not identical, the DRIVE dataset provides public CFIs that are generally similar to the ones provided in the UKBB. Hence, these CFIs were used to recompute these phenotypes and serve as a proxy for the performance of the methods in other datasets with the OD on one side.

For the number of bifurcations, the ground truth for these images was already available (http://www.retinacheck.org/download-iostar-retinal-vessel-segmentation-dataset, [26]), while validation by an ophthalmologist was required for the temporal angles.

#### 6.1 DRIVE temporal angles

For the temporal angles, both the arteries and the veins were analyzed separately. One image was removed because it did not have the OD on the side. The algorithm provides an angle when the majority votes agree; otherwise, it returns None. From the images with an angle, the performance was classified by an ophthalmologist as either 'good' or 'wrong'.

For the arteries, 26 images were classified as 'good' (0.87%), and 4 as 'wrong' (0.13%). Meanwhile, for the veins, 25 images were classified as 'good' (0.71%), and 10 as 'wrong' (0.29%).

The source of errors was analyzed. When there was a discontinuity in the vasculature segmentation, the algorithm detected a branch instead of the main vessels, and this was more frequent in veins. Additionally, in some cases, an anatomical variant with 2 main vessels was found, this was specially accused for veins. The complete table with the ophthalmologist comments can be found "Data DRIVE temporal angles".

#### 6.2 DRIVE number of bifurcations

Our method for counting bifurcations underestimated their number compared to manual annotations given in the DRIVE dataset. To investigate the relationship between the ground truth bifurcations and the segments detected as vasculature using LWNET, we plotted a validation analysis (Fig. 9), and both measurements exhibit a correlation of 0.65.

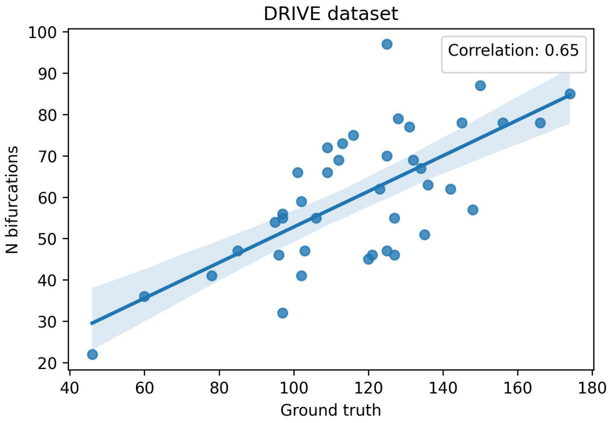


Figure 9: DRIVE ground truth: The x-axis represents the number of bifurcations, in the ground truth, that are over or close to the LWNET-detected vasculature segments, while the y-axis displays the number of bifurcations per CFI detected using our algorithm.

#### 7 Baseline characteristics

Main characterization of the UKBB subjects analyzed. As can be seen in the histograms (Fig. 6) some IDPs were possible to measure for most of the subjects (like the diameters, tortuosity, or vascular density), while IDPs that required the OD position tended to be measured for fewer samples due to issues in detecting it. However, the percentages remain similar. For two main IDPs, one representative of the first group ('A median diameter') and other of the other ('A temporal angle'), the baseline information, vascular and general diseases and ocular diseases can be found in the following table 1 and 2:

| | **A median diameter** | | | **A temporal angle** | | |
|---|---|---|---|---|---|---|
| **Parameters** | **Count** | **Mean** | **Std** | **Count** | **Mean** | **Std** |
| **Age Recruitment** | 68 579 | 56.34 | 8.07 | 55 329 | 55.73 | 8.1 |
| **Sex** | 68 579 | 0.46 | 0.5 | 55 329 | 0.46 | 0.5 |
| **Age Current Smoker** | 4 113 | 18.03 | 6.59 | 3 284 | 18.11 | 6.73 |
| **BMI** | 68 307 | 27.23 | 4.72 | 55 100 | 27.15 | 4.68 |
| **Age Diabetes** | 3 300 | 50.42 | 14.86 | 2 401 | 50.21 | 14.68 |
| **Age Angina** | 1 826 | 50.09 | 15.58 | 1 383 | 49.96 | 15.46 |
| **Age Heart Attack** | 1 358 | 52.03 | 11.67 | 1 018 | 51.51 | 11.76 |
| **Age DVT** | 1 303 | 41.45 | 16.7 | 1 020 | 40.78 | 16.76 |
| **Age Stroke** | 966 | 51.66 | 13.78 | 707 | 50.97 | 13.95 |
| **Age Pulmonary Embolism** | 514 | 44.81 | 14.98 | 407 | 44.21 | 14.61 |
| **Age Death** | 3 215 | 69.15 | 7.01 | 2 403 | 68.76 | 7.2 |
| **Age Glaucoma** | 1 179 | 51.82 | 15.22 | 845 | 51.97 | 14.42 |
| **Age Cataract** | 3 439 | 56.96 | 13.96 | 2 433 | 56.87 | 13.73 |
| **Eye Diabetes** | 634 | 2.53 | 0.77 | 435 | 2.54 | 0.77 |
| **Age Other Serious Eye Condition** | 1 250 | 44.11 | 18.93 | 933 | 44.85 | 17.97 |

Table 1: UKBB Summary of Baseline Characteristics, Disease Counts, Mean Values, and Standard Deviations for Various Parameters, including Age, Sex, BMI, and specific medical conditions (diabetes, angina, heart attack, DVT, stroke, pulmonary embolism, death, glaucoma, cataract, eye-diabetes, and other serious eye conditions) in the study population.

| IDP | N phenotypic | N GWAS |
|---|---|---|
| A temporal angle | 55 329 | 54 949 |
| V temporal angle | 57 875 | 57 486 |
| A tortuosity | 68 576 | 68 088 |
| V tortuosity | 68 499 | 68 012 |
| ratio tortuosity | 68 480 | 67 992 |
| A central retinal eq | 65 472 | 65 015 |
| V central retinal eq | 65 836 | 65 374 |
| ratio central retinal eq | 64 884 | 64 431 |
| A std diameter | 68 483 | 67 993 |
| V std diameter | 68 484 | 67 993 |
| bifurcations | 68 240 | 67 751 |
| A vascular density | 68 740 | 68 249 |
| V vascular density | 68 740 | 68 249 |
| ratio vascular density | 68 420 | 68 043 |
| A median diameter | 68 579 | 68 088 |
| V median diameter | 68 507 | 68 016 |
| ratio median diameter | 68 468 | 67 984 |

Table 2: Sample size per IDP. GWAS sample sizes are slightly smaller due to missing covariates, which we did not impute.

### 8 Covariate effects

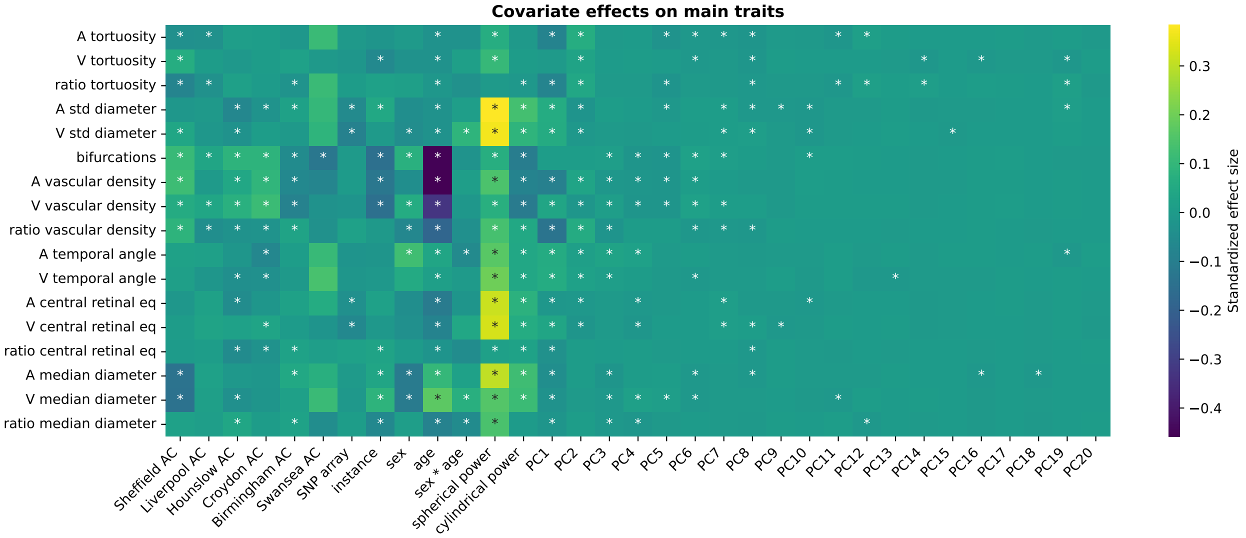


Figure 10: Standardized effects of considered covariates on main retinal IDPs. Categorical confounders: Assessment centers (ACs) where CFIs were taken (pairwise batch effects compared to Cheadle repeat assessment center), SNP arrays (UK BiLEVE Axiom array or Biobank Axiom array), instances (0 or 1). Numerical covariates: sex, age, sex * age, spherical power, cylindrical power, genetic PCs 1-20. Phenotypes and numerical covariates were z-scored. Squared covariates of age, sex*age, spherical power, and cylindrical power (which are used in the main analysis) were removed from this visualization model to avoid co-linearity in the design matrix, which leads to distorted p-value estimates. Asterisk: P-value significance threshold of $\alpha$=0.05 was not adjusted for multiple testing.

### 9 Replication of previously identified associations of retinal vascular phenotypes with diseases

In this section we compared our phenotypic associations between IDPs and diseases provided in the main text, with previous literature.

We replicated the negative association between hypertension and the diameter of the arteries (‘A median diameter’, ‘A central retinal eq’) [27, 28, 29, 30, 31, 32], as well as with the ratio between the diameters of arteries and veins (‘ratio median diameter’, and ‘ratio central retinal eq’) [33, 34, 35]. On the contrary, this association was not significant for the diameters or the veins (‘V median diameter’, and ‘V central retinal eq’, this second one with less evidence) [36, 28, 29, 31, 13, 14, 15, 16, 17, 18]. Additionally, we also replicated the positive association of hypertension with vascular tortuosity in both veins and arteries [23, 37, 1], although when considering SBD and DBP separately, only venular tortuosity was significantly associated.

Similarly, we replicated the negative association between high blood pressure and the vascular density of arteries (‘A vascular density’), the ratio between the vascular density of arteries and veins (‘ratio vascular density'), and the number of bifurcations ('bifurcations'). On the contrary, these associations were not confirmed in the case of vascular density or the veins ('V vascular density') [24, 21, 38].

Regarding age at death, we replicated the negative associations with vascular density [24] in the Cox model, while not in the logistic regression. However, we found no significant association between stroke and arterial vessels' diameter despite this having been reported in earlier studies [39, 40, 28].

For type-2 diabetes, we replicated a positive significant association with the diameter of arteries ('A median diameter', and 'A central retinal eq') and the median diameters of veins ('V median diameter', this last one with less evidence), but not for the central retinal equivalent of the veins ('V central retinal eq') [41, 42, 43]. The arterial median diameters (but not the venous) association held true also in the case of diabetic retinopathy, as suggested by previous studies with smaller sample sizes [44].

Additionally, we replicate the positive association between angina and tortuosity [1], particularly we only found this association for veins ('V tortuosity').

Regarding the association between IDPs and ophthalmological diseases, we replicated negative associations between glaucoma and cataract and vascular density, both arteries and veins, and the number of bifurcations ('A vascular density', 'V vascular density', and 'bifurcations') [24, 45], although we did not find this in the case of the ratio of vascular densities. We found that the number of bifurcations followed these same association patterns.

Myopia and presbyopia show discordant association profiles. We replicated associations between myopia was associated with temporal angles of arteries and veins [5]. Additionally, hypermetropia showed associations with the median diameter of vessels. No significant associations were found in the case of astigmatism, despite reports of strong genetic determinants for this phenotype [46].

### 10 Manhattan and Quantile-Quantile plots

Manhattan and Quantile-Quantile (QQ) plots of the 17 main IDPs used in the main paper (Fig. 11 and 12, respectively) show that the different IDPs exhibit heterogeneous patterns, indicating variations in their genetic associations. The IDPs with the highest number of significant SNPs are primarily associated with tortuosity. Conversely, the IDPs related to median diameters exhibit the lowest number of significant SNPs. Furthermore, notable differences in patterns emerge when comparing the genetic associations between arteries and veins within the same IDP. These variations imply distinct genetic influences on arterial and venous vascular phenotypes, highlighting the importance of considering vessel type specificity in genetic studies of retinal IDPs.

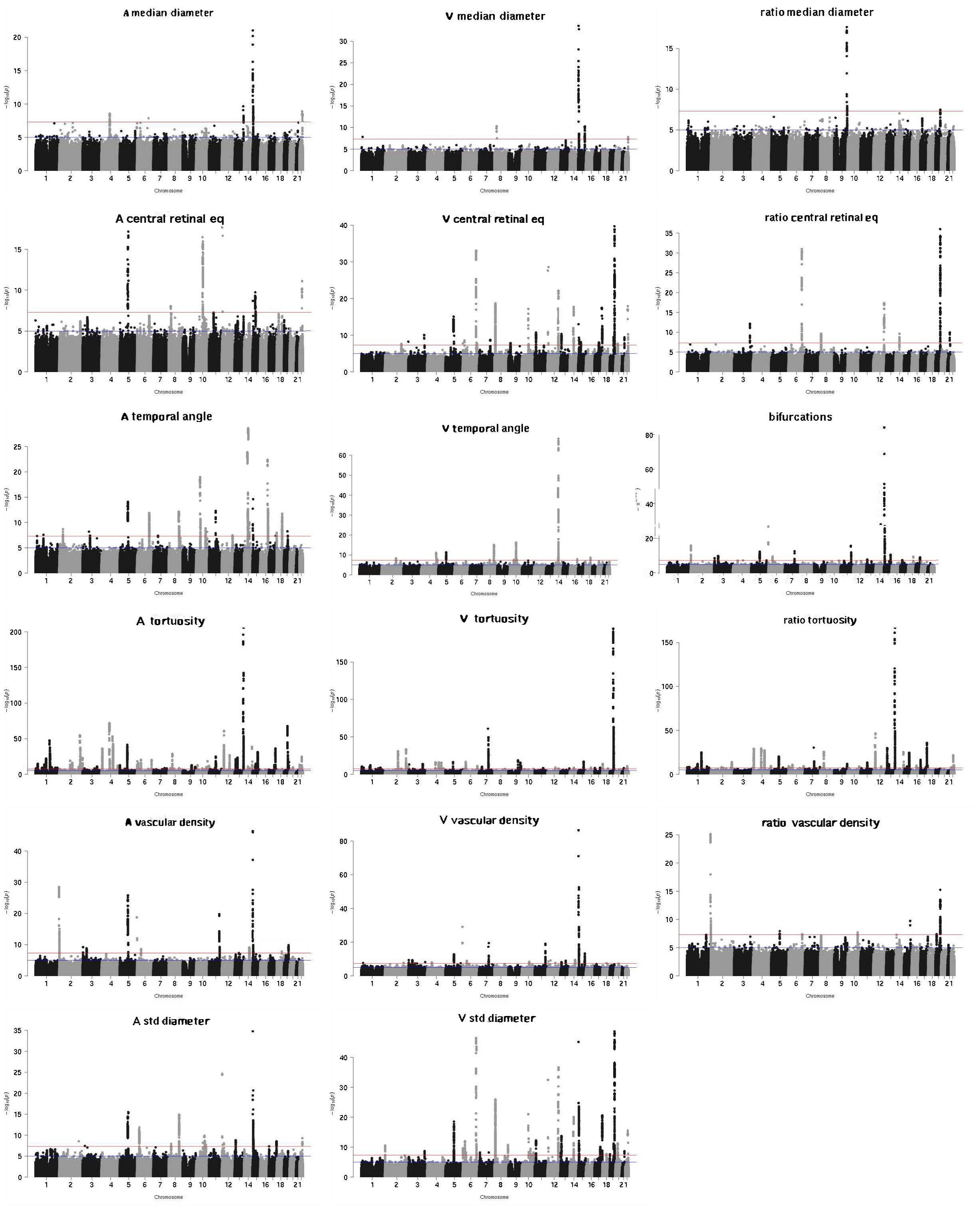


Figure 11: Manhattan plots of the 17 main IDPs used in the main paper from the UKBB. Each subfigure represents a different IDP. The x-axis displays the positions across the 22 Chromosomes, while the y-axis represents the -log10(p) associated with each SNP. The red line indicates the Bonferroni threshold, above which the dots are considered significantly associated.

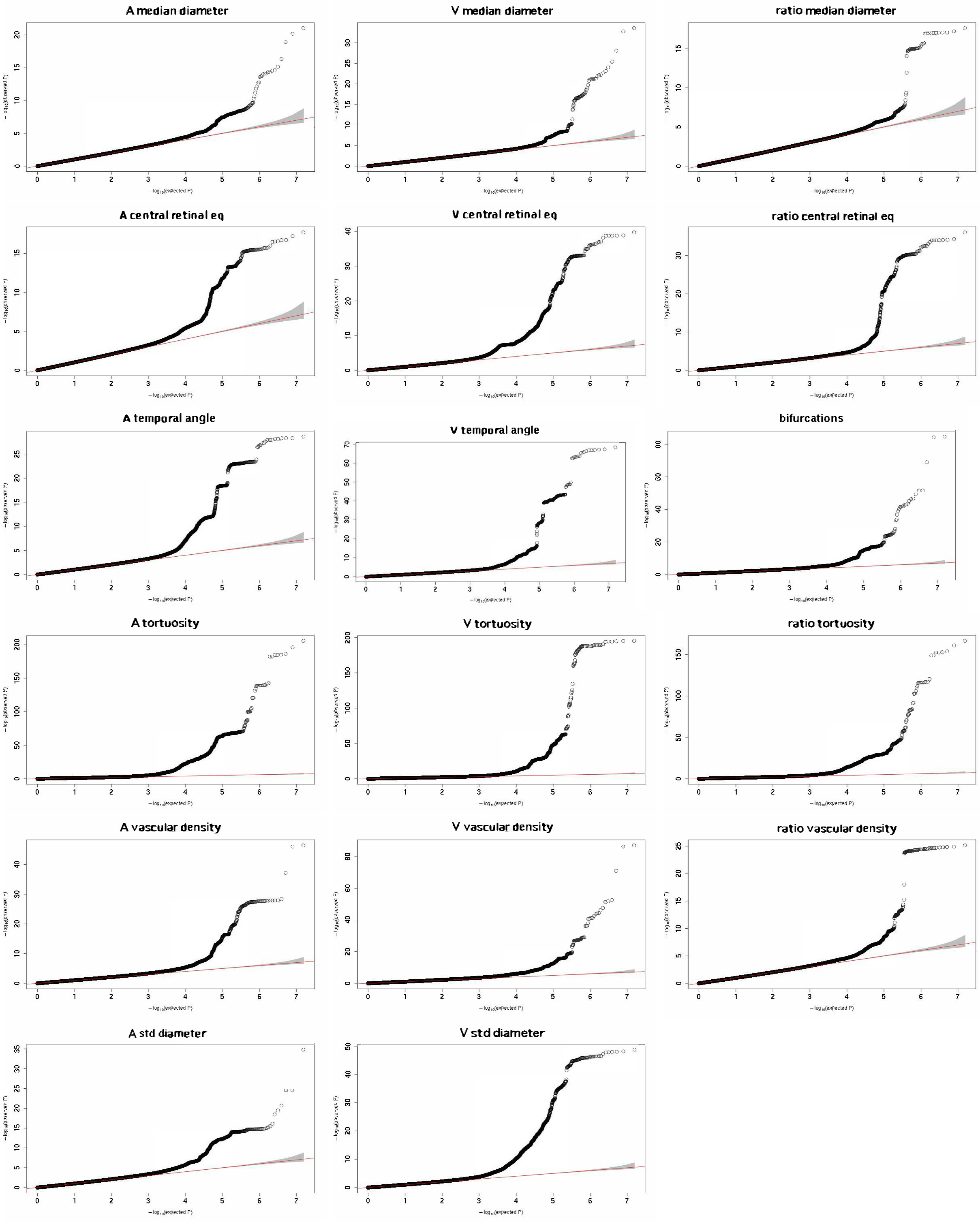


Figure 12: QQ plots of the 17 main IDPs used in the main paper from the UKBB. Each subfigure represents a different IDP. The x-axis represents expected quantiles, while the y-axis represents observed quantiles. The points on the plot compare the observed quantiles of the IDPs to the expected quantiles based on a theoretical distribution. The alignment between observed and expected quantiles indicates the similarity between the IDPs' distribution and the theoretical distribution.

### 11 Comparison with previous GWAS

To assess the consistency of our measurements for the retinal vascular IDPs that have been previously computed in GWAS, namely tortuosity, fractal dimension, vascular density, and diameter, we aim to compare our results with the existing findings.

Table 3 provides a summary of the associated SNPs with these various retina vascular IDPs. This table includes information on the IDP, the sample size and ancestry of the discovery and replication cohorts, the count of SNPs associated with each phenotype, some of the genes that are implicated in these associations, and the corresponding references for further details. By examining this table, researchers and readers can gain insights into the specific SNPs, associated genes, and the existing literature on the genetic basis of various retina vascular IDPs.

With the exception of one study [1], the selection of genes associated with IDPs relied solely on the strength of the association of individual SNPs. However, this method lacks robustness in identifying significant genes, as it only considers individual contributions with strong associations.

| IDP | Discovery | Replication | Association count (SNPs) | Genes associated | Reference |
|---|---|---|---|---|---|
| | Sample size and ancestry | | | | |
| | | | **Diameter vessel based:** | | |
| **CRAE** | 24 275* | 5 472 Europ | 1 | *OCA2* | [18] |
| | 18 722 Europ | 3 939 Europ | 1 | *MEF2C* | [17] |
| **CRVE** | 24 275* | 5 472 Europ | 4 | *TEAD1*, *GNB3*, *TSPAN10*, *OCA2* | [18] |
| | 15 358 Europ | 6 652 Europ | 4 | *RASIP1*, *VTA1*, *ATXN2*, *MEF2C* | [19] |
| **AVR** | 3 094 Europ | - | 1 | *SIX2/LINC01121* | [47] |
| **Width$_A$** | 52 798 Europ | - | 2 | *GNB3*, *CLUL1* | [48] |
| **Width$_V$** | 52 798 Europ | - | 11 | *CFHR4*, *MEF2C*, *NMBR*, etc | [48] [48] |
| | | | **Tortuosity vessel based:** | | |
| **Max($\tau_A$)** | 3 094 Europ | - | 2 | *LINC00917*, *COL4A2* | [47] |
| **Max($\tau_V$)** | 3 094 Europ | - | 1 | *AJAP1* | [47] |
| **Mean($\tau_A$)** | 3 094 Europ | 1 413 Europ | 2 | *COL4A2* | [47] |
| **Median($\tau_A$)** | 63 662 Europ | 911 Europ | 116 | *ACTN4*,*COLCA2*, *TNS*, etc | [1] |
| **Mean($\tau_V$)** | 3 094 Europ | 1 413 Europ | 1 | *ACTN4/CAPN12* | [47] |
| **Median($\tau_V$)** | 63 662 Europ | 911 Europ | 63 | *ACTN4*,*TNS1*, *CAPN12*, etc | [1] |
| **Median($\tau$)** | 63 662 Europ | 911 Europ | 125 | *TNS1*, *COL4A2*, *ACTN4*, etc | [1] |
| $\tau_A$ | 52 798 Europ | - | 89 | *COL4A2*, etc | [48] |
| $\tau_V$ | 52 798 Europ | - | 17 | *ACTN4*,etc | [48] |
| | | | **Vascular branching complexity based:** | | |
| **FD** | 54 813 Europ | - | 7 | *OCA2*, *COLCA1*, *SLC45A2*, etc | [24] |
| | | | **Vascularisation density based:** | | |
| $\boldsymbol{\rho_v}$ | 54 813 Europ | - | 13 | *MEF2C*, *GNB3*, *COLCA1*, etc | [24] |
| | | | **Auto encoder endophenotypes:** | | |
| **Endo$_{IDPs}$** | 65 629 Europ | - | 1 214 | *FLT1*, *EPHB4*, *WNT7B*, etc | [49] |

Table 3: Summary of previous retinal vascular IDPs GWAS: SNPs association count reported is this resulting after LD correction. 24 275*: 14 740 European, 1 077 Hispanic or Latin American, 3 345 African American or Afro-Caribbean, 5 113 South Asian, South East Asian, East Asian. Europ=European. IDPs names: Central retinal equivalent (CRE), Diameter Artery Vein ratio (AVR), Tortuosity ($\tau$), Fractal dimension (FD), Vascular density $\boldsymbol{\rho_v}$, latent variables from the autoencoder (Endo$_{IDPs}$).

Based on our results from the main paper, "Data Significant genes per IDP", and the previous studies: The genes *OCA2*, and *MEF2C* associated with 'A central retinal eq' (CRAE), were not found as significant in our analysis. For the 'V central retinal eq' (CRVE), the genes *TEAD1*, *GNB3*, *RASIP1*, *VTA1*, and *ATXN2* were significant in our analysis, while not *TSPAN10*, *OCA2*, *MEF2C*. And regarding the genes associated with the ratio *SIX2/LINC01121*, none of them were found to be significantly associated with any IDPs.
Regarding the $Width_A$, *GNB3* is significantly associated with 'V std diameter'. While for $Width_V$, *MEF2C* with 'V central retinal eq', *NMBR* with 'ratio central retinal eq', 'V std diameter', and 'V central retinal eq', *XKR6* with 'V std diameter', and 'V central retinal eq', *MYPN* with 'V temporal angle' and 'A central retinal eq', 'FLT1' with 'ratio tortuosity', 'A std diameter', 'V std diameter', 'A tortuosity', and 'V central retinal eq', *WDR72* with 'A central retinal eq', *FUT2* with 'ratio central retinal eq', 'V std diameter', 'V central retinal eq', and 'ratio vascular density'.
Regarding the genes associated with Max($\tau_A$) and Max($\tau_V$), only *COL4A2* was significant in our IDPs, particularly with 'A tortuosity' and 'ratio tortuosity'. Regarding the genes associated with Mean($\tau_A$) and Mean($\tau_V$), *ACTN4* and *CAPN12* are both associated with 'V tortuosity', 'A tortuostity', 'V std diameter', and 'V central retinal eq', and *COL4A2* is associated with 'A tortuosity', and 'ratio tortuosity'.
Regarding Median($\tau$), Median($\tau_A$), Median($\tau_V$), since the dataset is the same (except QC and segmentation) and most of the methodology is similar we can see that most are the same for both, arteries and veins respectively with [1]. Additionally, [1] also explored additional tortuosity definitions based on curvature, two of which were analyzed when selecting the main IDPs, their additional parameters were not explored further than their association with diseases and heritability, but we can see that for $\tau_2$ heritability values are slightly smaller in our current analysis $h^2 \pm$std$= 0.07 \pm 0.010$ than the ones reported $h^2 = 0.11 \pm 0.011$, while for $\tau_4$, we get almost twice the previous value, $h^2 = 0.22 \pm 0.023$ instead of $h^2 = 0.12 \pm 0.011$. This could be attributed to the definition of tortuosity based on curvature is highly sensitive to the segmentation method used, and improving it also improves the accuracy of the tortuosity measurement.
Finally, regarding the endophenotypes, *SH3YL1* is associated with 'ratio vascular density', 'A vascular density', 'bifurcations', and 'V std diameter'. *CTNNB1* with 'A vascular density', 'bifurcations', and 'V vascular density'. *FGFR3* with 'ratio tortuosity'. *GABRB1*, *EYS*, and *ZKSCAN1* with 'V vascular density'. *IRF4* with 'bifurcations', and 'V vascular density'. *FOXP4* with 'A vascular density', 'bifurcations', and 'V vascular density'. *PDE3A* with 'A tortuosity', 'ratio tortuosity', and 'A std diameter'. *FLT1* with 'A tortuosity', 'ratio tortuosity', 'A std diameter', 'V std diameter', and 'V central retinal eq'. *HERC2* with 'V std diameter', 'A vascular density', 'bifurcations', 'V vascular density', 'A median diameter', and 'V median diameter'. *SLC47A1* with 'bifurcations', and 'V vascular density'. *PDE6G* with 'A std diameter', 'V std diameter', and 'V central retinal eq'. And *WNT7B* with 'V std diameter', and 'V central retinal eq'.

### 12 Genetics associations between vascular IDPs and binary diseases

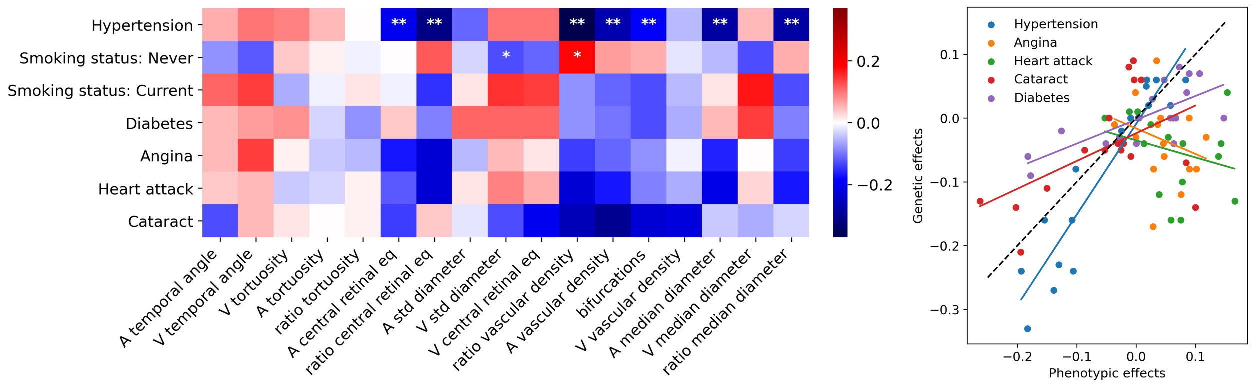


(a) (b)

Figure 13: **a)** Genetic correlation between IDPs and binary diseases, computed using LDSR [50] in the UKBB. The color indicates the genetic correlation coefficient and the asterisks indicate the level of statistical significance ($* : p < 0.05/N_{test}$, $** : p < 0.001/N_{test}$, being $N_{test} = N_{IDPs} \times N_{LinearDiseases}$). **b)** Correlation between phenotypic and genetic correlations of IDPs with binary diseases, in the UKBB.

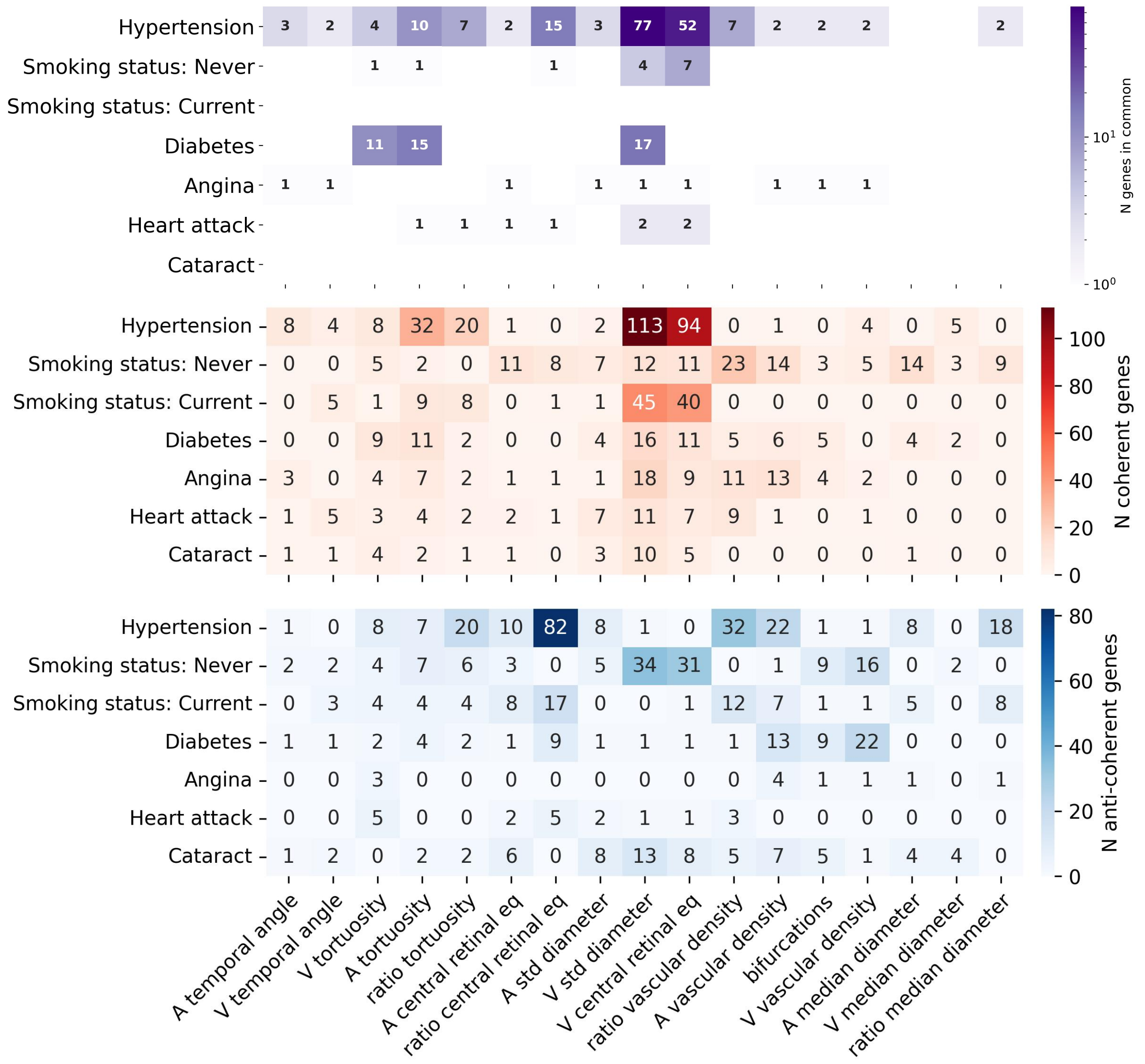


Figure 14: **a)** Gene-scoring plain intersection showing genes in common between IDPs and binary diseases, in the UKBB. Each cell shows the number of intersected genes in phenotype pairs. **b)** and **c)** Cross-phenotype coherence analysis showing the number of coherent **b)** and anti-coherent **c)** genes between phenotype pairs. Summary statistics for cases control diseases were obtained from http://www.nealelab.is/uk-biobank, and significant genes were computed using *PascalX* [51].

### 13 MR analysis between vascular IDPs and binary diseases

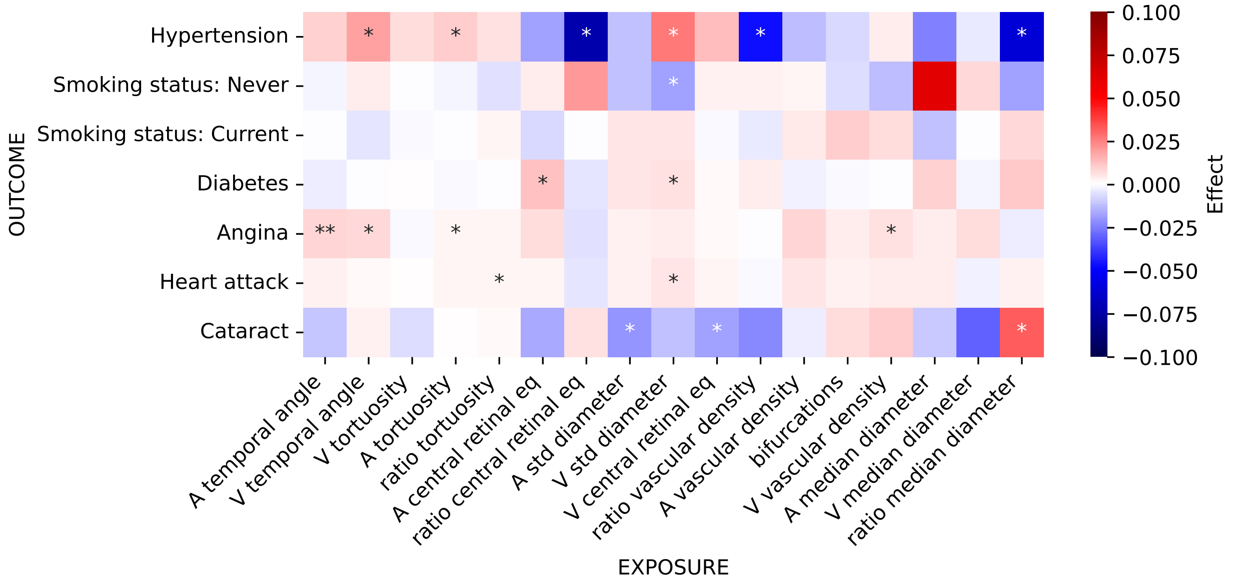


(a)

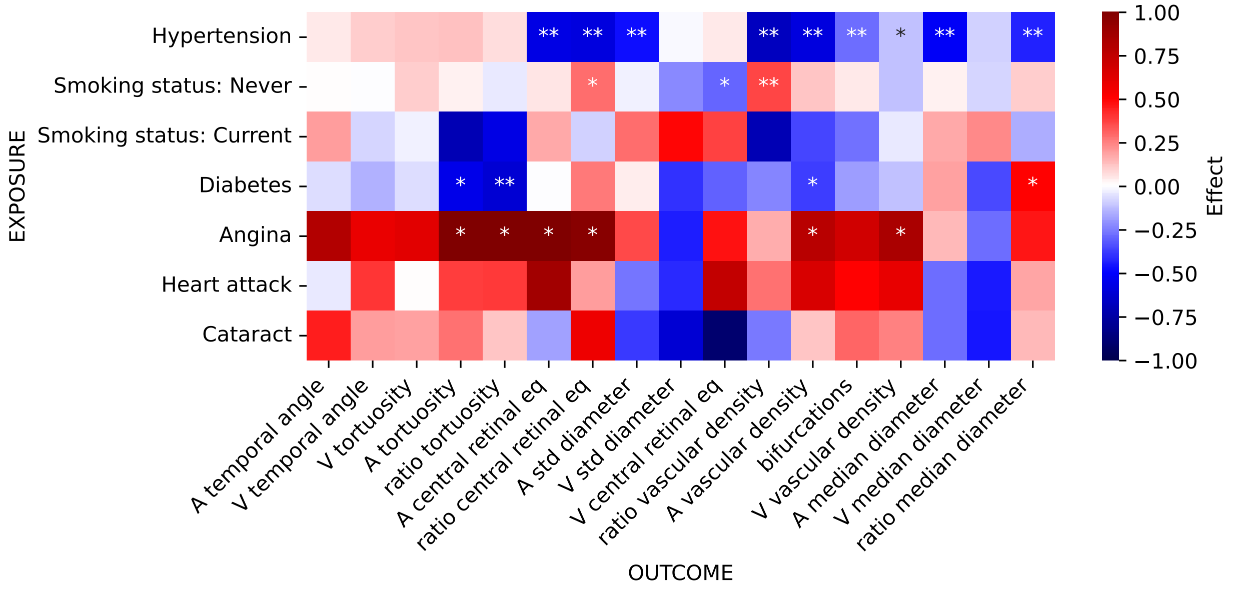


(b)

Figure 15: **a)** Causal effect estimates with IDPs as exposures and binary diseases as outcomes, in the UKBB. **b)** Causal effect estimates with binary diseases as exposures and IDPs as outcomes, in the UKBB. The color indicates the causal effect estimates based on the inverse variance-weighted MR method. The level of statistical significance is indicated with a single asterisk for nominal significance without correction for multiple testing ($* : p_{uncorrected} < 0.05$) and two asterisks for a false discovery rate (FDR) ($** : p_{FDR} < 0.05$).

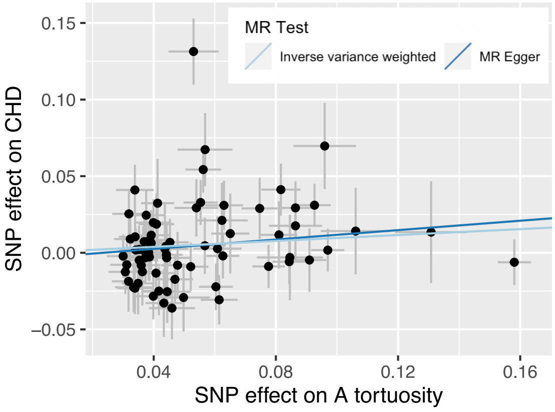


| outcome | exposure | nsnp | b | se | pval |
|---|---|---|---|---|---|
| CHD | bifurcations | 12 | -0.073198095 | 0.161850288 | 0.651083097 |
| CHD | A std diameter | 10 | 0.017461403 | 0.263426777 | 0.947150444 |
| CHD | V std diameter | 24 | 0.195441647 | 0.122132217 | 0.109543921 |
| CHD | A central retinal eq | 12 | 0.102256346 | 0.180455163 | 0.570946621 |
| CHD | V central retinal eq | 21 | 0.155870693 | 0.148389256 | 0.293526137 |
| CHD | A temporal angle | 18 | 0.088372474 | 0.110059884 | 0.422004211 |
| CHD | V temporal angle | 11 | 0.166277063 | 0.13203337 | 0.207901648 |
| CHD | A median diameter | 5 | 0.131777368 | 0.247331782 | 0.594174847 |
| CHD | V median diameter | 4 | 0.176738774 | 0.245488881 | 0.471558155 |
| CHD | ratio tortuosity | 43 | 0.074152477 | 0.064465716 | 0.250035892 |
| CHD | ratio median diameter | 4 | 0.627877102 | 0.631479388 | 0.320079035 |
| CHD | ratio central retinal eq | 11 | -0.299970255 | 0.160835531 | 0.062170955 |
| CHD | ratio vascular density | 13 | -0.143407548 | 0.159432831 | 0.368394022 |
| CHD | A tortuosity | 72 | 0.09641616 | 0.048477503 | 0.046713942 |
| CHD | V tortuosity | 35 | 0.052211027 | 0.078783138 | 0.507510987 |
| CHD | A vascular density | 18 | 0.063858105 | 0.150230195 | 0.67078683 |
| CHD | V vascular density | 13 | 0.052665908 | 0.156019999 | 0.73569577 |

Figure 16: Results of MR analyses for arteriolar tortuosity as a causal influence on coronary heart disease (CHD) in the UKBB. **Left:** We were able to confirm this causal link of arterial tortuosity on CHD [48]. Fit lines correspond to inverse-variance-weighted (IVW) and Egger methods. **Right:** Effects from IVW for all IDPs are reported. No other retinal IDP was found to be causal.

### 14 LDSR genetic correlation against PascalX

The genetic correlation between IDP pairs and between IDPs and risk factors was computed at the SNP level using LDSR and at the gene level using PascalX cross-GWAS coherence tests. Generally, phenotype pairs with positive SNP-wise genetic correlation tended to have more coherent than anticoherent gene signals, while pairs with negative SNP-wise correlation had more anticoherent gene signals (Fig. 17 and 18). This was especially true when gene-wise signals were 100% coherent or 100% anticoherent. For pairs that shared a mixture of coherent and anticoherent genes, SNP-wise correlation did not always follow the same direction as gene signals, suggesting that more nuanced and interpretable results might be obtained by aggregating SNPs onto genes.

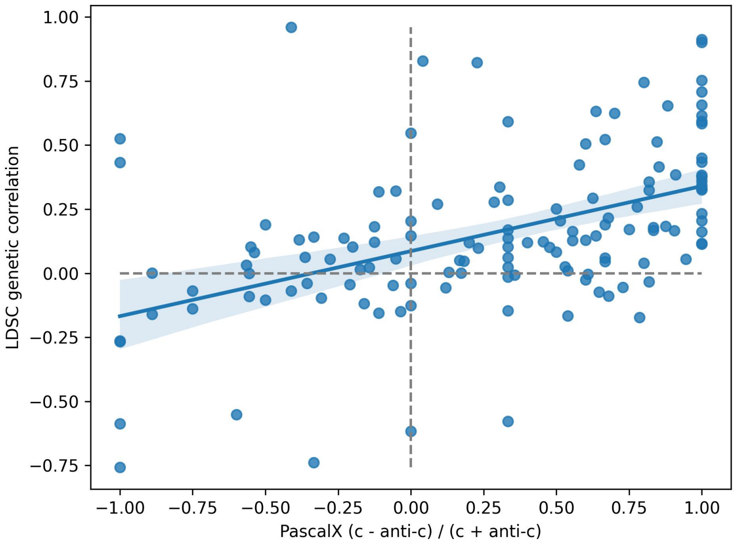


Figure 17: IDP-IDP pairs: LDSR genetic correlation plotted against PascalX normalized coherence (number of coherent-anticoherent genes)/(number of coherent+anticoherent genes). Each dot represents a phenotype pair. The regression line is plotted with a 95% confidence interval. 'LDSR': Linkage Disequilibrium Score Regression, 'c': coherent, 'anti-c': anticoherent.

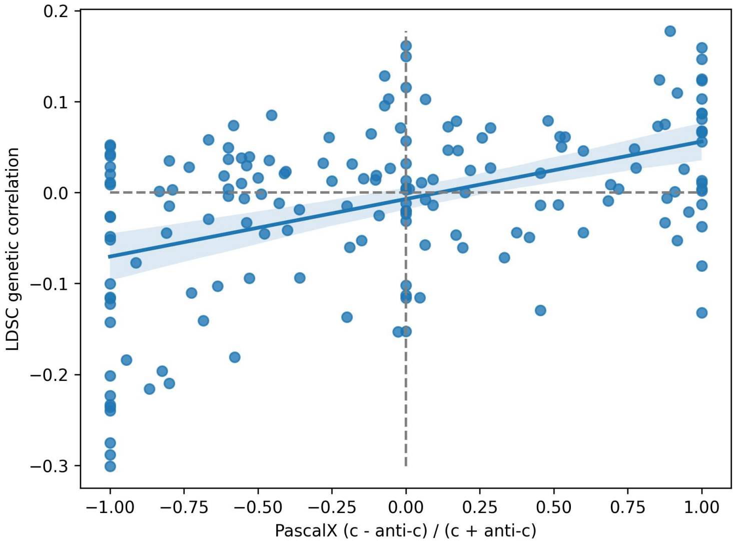


Figure 18: IDP-risk factor pairs: LDSR genetic correlation plotted against PascalX normalized coherence (number of coherent-anticoherent genes)/(number of coherent+anticoherent genes). Each dot represents a phenotype pair. The regression line is plotted with a 95% confidence interval. 'LDSR': Linkage Disequilibrium Score Regression, 'c': coherent, 'anti-c': anticoherent.

### 15 Pathway analyses

We analyzed the significant pathways (sets of genes) that are significantly associated with each retina vascular IDP. In this case, only the tortuosity of the arteries has more than 40 pathways significantly associated, right Fig 19. Outside this figure's diagonal, we see that no pathways are common to pairs of IDP. Again, there are no common pathways for all of the IDPs, and from Fig. 19, we can see fewer pathways common to multiple IDPs. The most frequent pathways are not only exclusive to the retina, such as 'Abnormal retinal morphology', but also relate to other organs, such as the midbrain 'Midbrain neurotypes hendo', and the kidney 'Lake adult kidney C27 vascular smooth muscle cells and pericytes'.

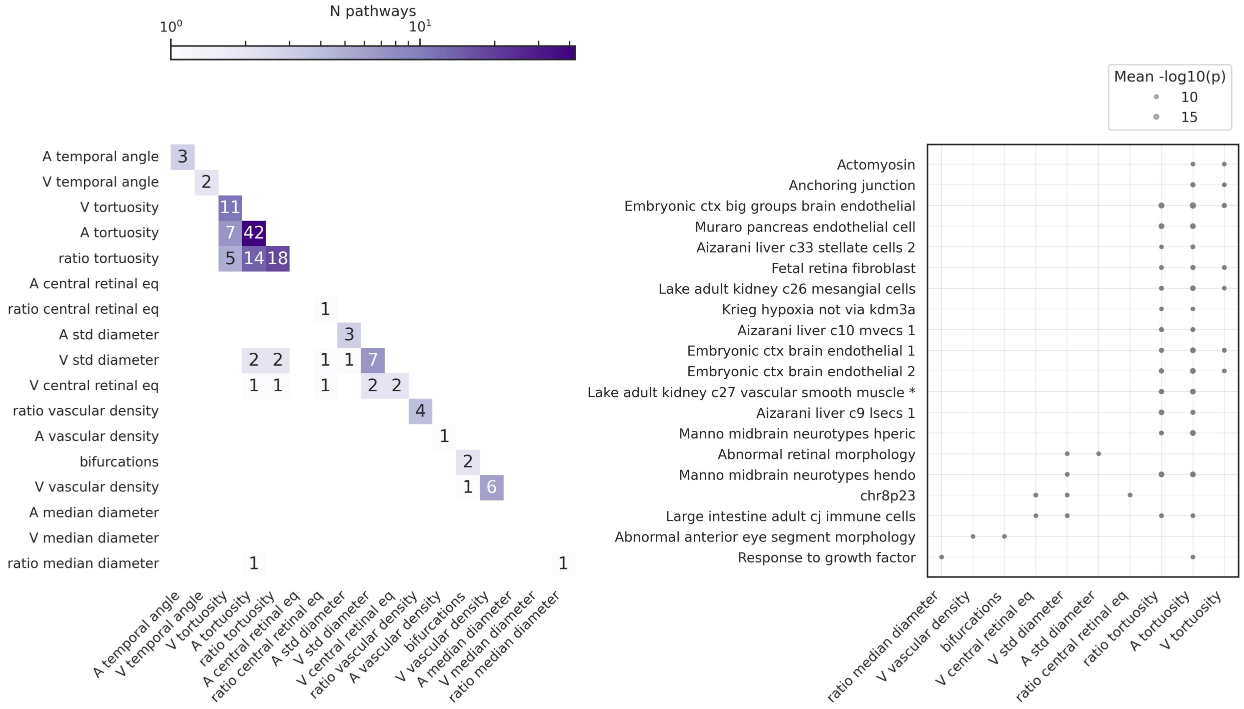


Figure 19: **a)** Shared pathways through the different IDPs, in the UKBB. The number of pathways significantly associated per IDP can be found in the diagonal, the upper diagonal contains the number of shared pathways of phenotype-pairs. **b)** Top most frequent pathways through the different IDPs. Dot sizes depend on the p-value associated [51].

### 16 Pathway analyses between vascular IDPs and diseases

The GWAS of the previously mentioned diseases were analyzed using *PascalX* to obtain the pathways significantly associated with each of these diseases, "Data Significant pathways per diseases". Subsequently, the intersection was performed between each of the pathways associated with these diseases as well as with the vascular retinal IDPs, resulting in the following figure 20, which shows for each disease-IDP pair the number of significant pathways they have in common.

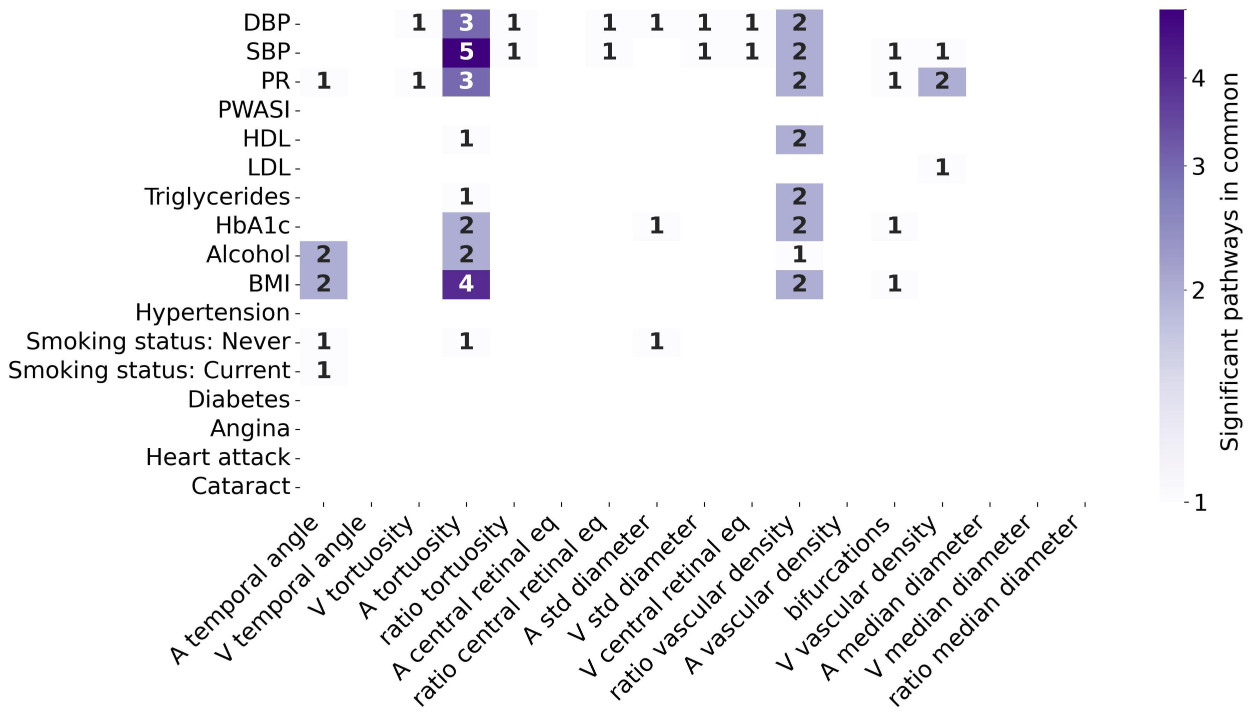


Figure 20: Intersection shared pathways through the different IDPs and the Neale ss diseases. The diseases are the same as those used for genetic correlation and gene analysis.

### 17 Replication

#### 17.1 The Rotterdam Study

The Rotterdam Study (RS) is a prospective population-based cohort study of people living in Ommoord, a district of the city of Rotterdam [52]. The RS consists of four cohorts, all of which were used in this replication. The first cohort (RS-I) started in 1991 and consisted of 7 983 participants $>$ 55 years (range 55.0 – 99.2, response rate of 78%). The second cohort (RS-II) started recruiting in 2 000 and 3 011 participants of $>$ 55 years were included (range 55.2 – 98.9, response rate of 67.3%). The third cohort (RS-III) also included people aged $>$ 45 years (range 45.7-90.1) and consisted of 3 932 participants (response rate 64.9%) starting from the year 2006. The fourth cohort (RS-IV) included people aged $>$ 40 years and consisted of 3 005 participants. Follow-up examinations were performed every 4 to 5 years. The RS is entered into the Netherlands National Trial Register (NTR) and the WHO International Clinical Trials Registry Platform (ICTRP) under shared catalog number NTR6831. All participants provided written informed consent following the Declaration of Helsinki to participate in the study and to have their information obtained from their treating physicians.

#### 17.2 CoLaus and OphtalmoLaus

The CoLaus study, initiated in 2003 in Lausanne, Switzerland, involves over 6 700 volunteers aged 35 to 75, accounting for about 10% of the city's population. Through periodic examinations, it aims to identify cardiovascular and psychiatric health correlations. OphtalmoLaus, a segment of CoLaus, delves into ocular health, studying eye disease prevalence and genetics. The effort has led to over 650 publications, merging insights on cardiovascular, psychiatric, and ocular health. In OphtalmoLaus, different types of eye images are acquired, including optical coherence tomography (OCT) cubes and lines, OCT-A, Iris pictures, and CFIs. These were acquired with Topcon 2000 or Topcon triton.

#### 17.3 Methodology

The overall steps of the replication in the RS were the following:

**Data selection** We used RS data from recent rounds (RS-I-4, RS-II-4, RS-III-1, RS-IV-1) due to the higher quality of the imaging and availability of more images for each visit/participant. We discarded images where the OD was near or out of the CFI boundaries since this often indicated a lack of visibility of the major vessels. This was done automatically by 1) detecting the CFI boundaries using an algorithm developed in house and empirically validated on RS data and 2) segmenting the OD using a DL segmentation model similarly developed in house and validated on RS CFIs. Images for which any pixel in the OD mask was under a small threshold (5px) distance to the fundus bounds were discarded.

**IDP extraction** The output of DL models trained on RS data for disc segmentation, fovea localisation, and artery-vein segmentation were used for feature extraction. The 17 phenotypes used in the main study were implemented in vascular analysis software developed in house. Figure 21 shows illustrations of some of the feature computations including the use of disc-centered images. The pseudo-code in Section 2 was adapted when necessary to work on these images.

**IDP outlier elimination** Per cohort, IDPs more than 10 standard deviations away from the mean were eliminated as outliers.

**Individual GWAS** For the initial GWAS analyses, we performed linear regressions using Plink2.0 [53], with the allele dosage values as the exposure and the IDP as the outcome. The included covariates were age, sex, spherical equivalent, imaging device (coded as dummy variables), and genomic PCs 1-10.

**Meta-GWAS** We performed an inverse variance weighted fixed-effect meta-analysis over the four RS cohorts, using METAL software [54]. P-values for the association results were calculated by using the z-statistic. The meta-analyzed significant hits were pruned using Plink2.0. Variants were considered independent if they were at least 500 kb apart and had an $R2 < 0.1$.

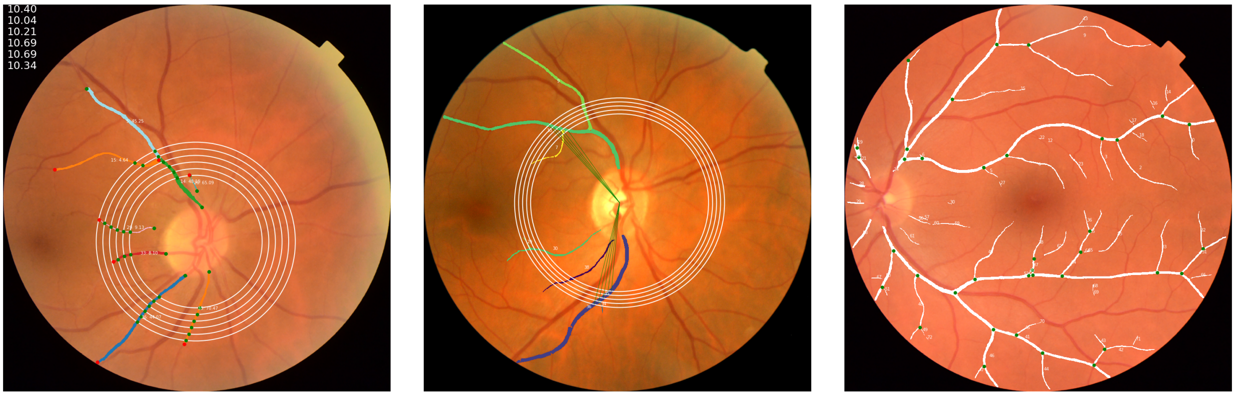


(a) V central retinal equivalent (b) V temporal angle (c) Number of bifurcations

Figure 21: Visualization of some IDPs on RS images.

Regarding OphtalmoLaus, the steps of the methodology replication were the same as in the UKBB, 1 for vessel segmentation and 2 for IDPs extraction and [24] for QC.
Some IDPs were possible to measure for most of the subjects (like the diameters, tortuosity, or vascular density), while phenotypes that used the OD as a reference tended to be measured for fewer samples due to issues in measuring it. However, the percentages remain similar. For two main IDPs, one representative of the first group ('A median diameter') and other of the other ('A temporal angle'), the baseline information can be found in the following table 4:

| | A median diameter | | | A temporal angle | | |
|---|---|---|---|---|---|---|
| **Parameters** | **Count** | **Mean** | **Std** | **Count** | **Mean** | **Std** |
| **Age Recruitment** | 1 426 | 64.16 | 9.15 | 1 018 | 62.94 | 8.64 |
| **Sex** | 1 426 | 0.54 | 0.50 | 1 018 | 0.55 | 0.50 |
| **BMI** | 1 424 | 25.02 | 3.92 | 1 016 | 24.82 | 3.83 |

Table 4: OphtalmoLaus Summary of Baseline Characteristics, Mean Values, and Standard Deviations for Age, Sex, and BMI.

#### 17.4 IDPs phenotypic replication

Figure 22 shows the distribution of IDPs in the RS. Comparison with Figure 6 reveals similar relative distributions of artery and vein IDPs to the UKBB. It is important to note that IDPs affected by the scale of the images such as median diameters, CREs, and vascular densities are not directly comparable. Differences in the shape of distributions such as temporal angles and vascular densities are likely due to the use of different segmentation methods, differences in image quality across datasets, and the differences in implementation mentioned in Section 17.3.

Additionally, figure 23 shows the association of IDPs with some risk factors and diseases in the RS.

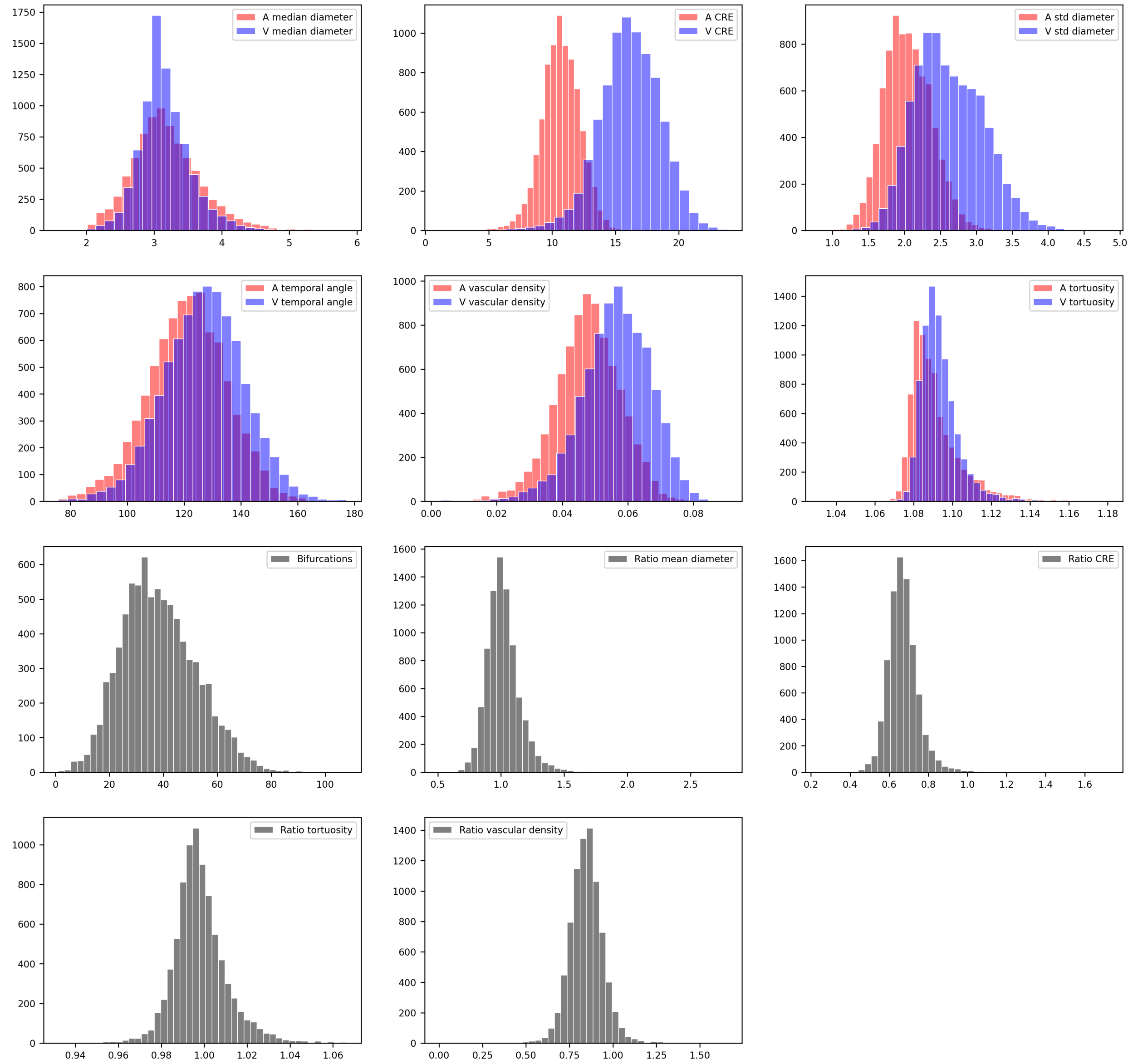


Figure 22: Phenotypic distributions in the RS. Raw distributions of the 17 main IDPs per subject. Outlier values were removed. This outlier removed was especially important for the ‘ratio vascular density’.

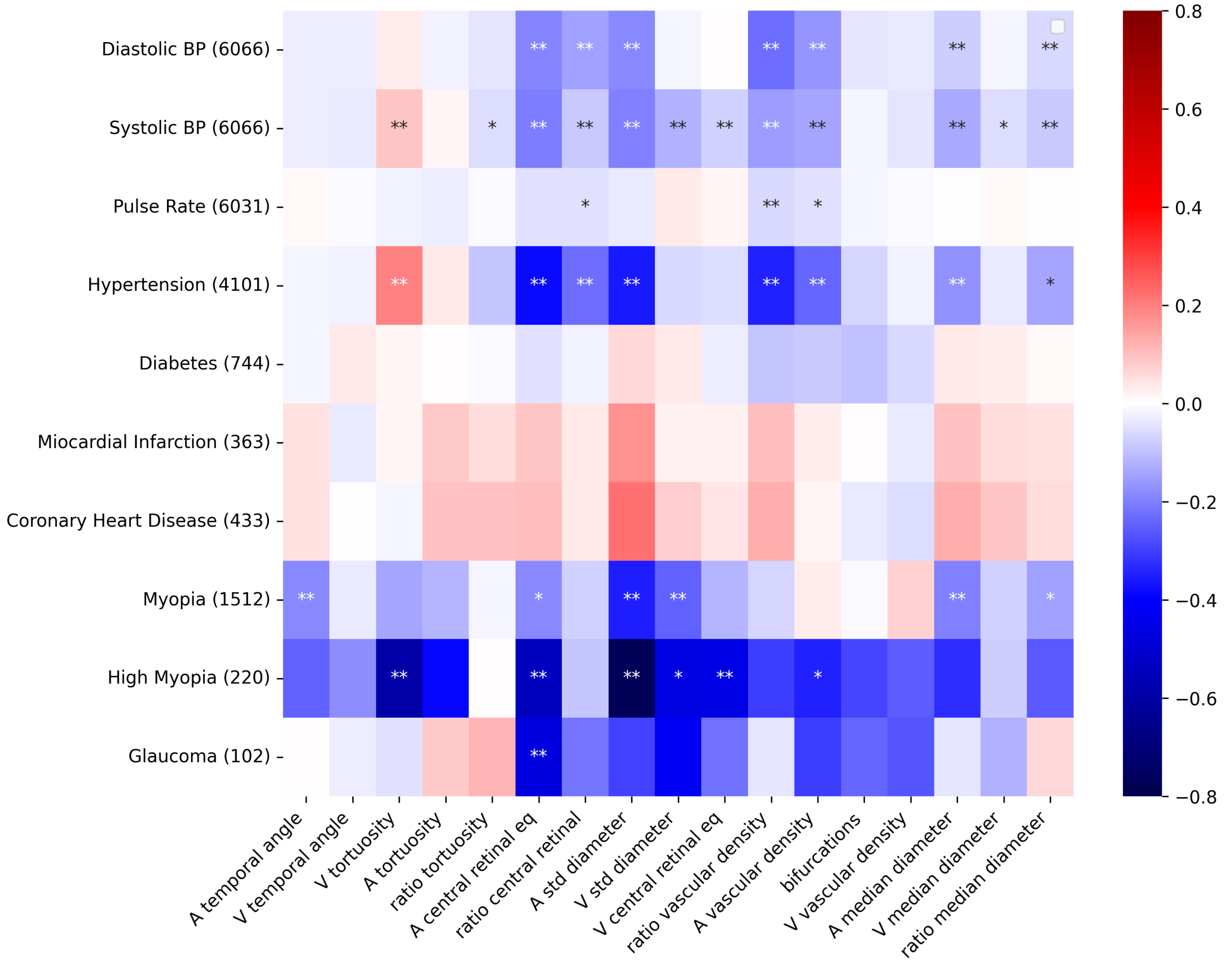


Figure 23: Phenotypic association of IDPs with diseases in the RS. The x-axis shows IDPs and the y-axis shows diseases. For continuous variables, the numbers in parentheses correspond to the number of subjects with available data, and for binary disease states to the number of cases. Linear (a) and logistic (b) regressions were used for continuous and binary disease states respectively. In all models, phenotypes were corrected for age, sex, eye geometry, and imaging device. The color indicates standardized effect sizes (Standardized Beta) for linear and logistic regressions. Asterisks indicate the level of statistical significance ($* : p < 0.05/Ntests$, $** : p < 0.001/Ntests$, where $Ntests = NIDPs \times Ndiseasetraits$, and $Ndiseasetraits$ is the number of diseases (traits) considered in each panel).

Similarly, figure 24 shows the distribution of IDPs in OphtalmoLaus. Comparison with Figure 6 reveals again very similar relative distributions of artery and vein IDPs.

Additionally, figure 25 shows the association of IDPs with risk factors and diseases in OphtalmoLaus.

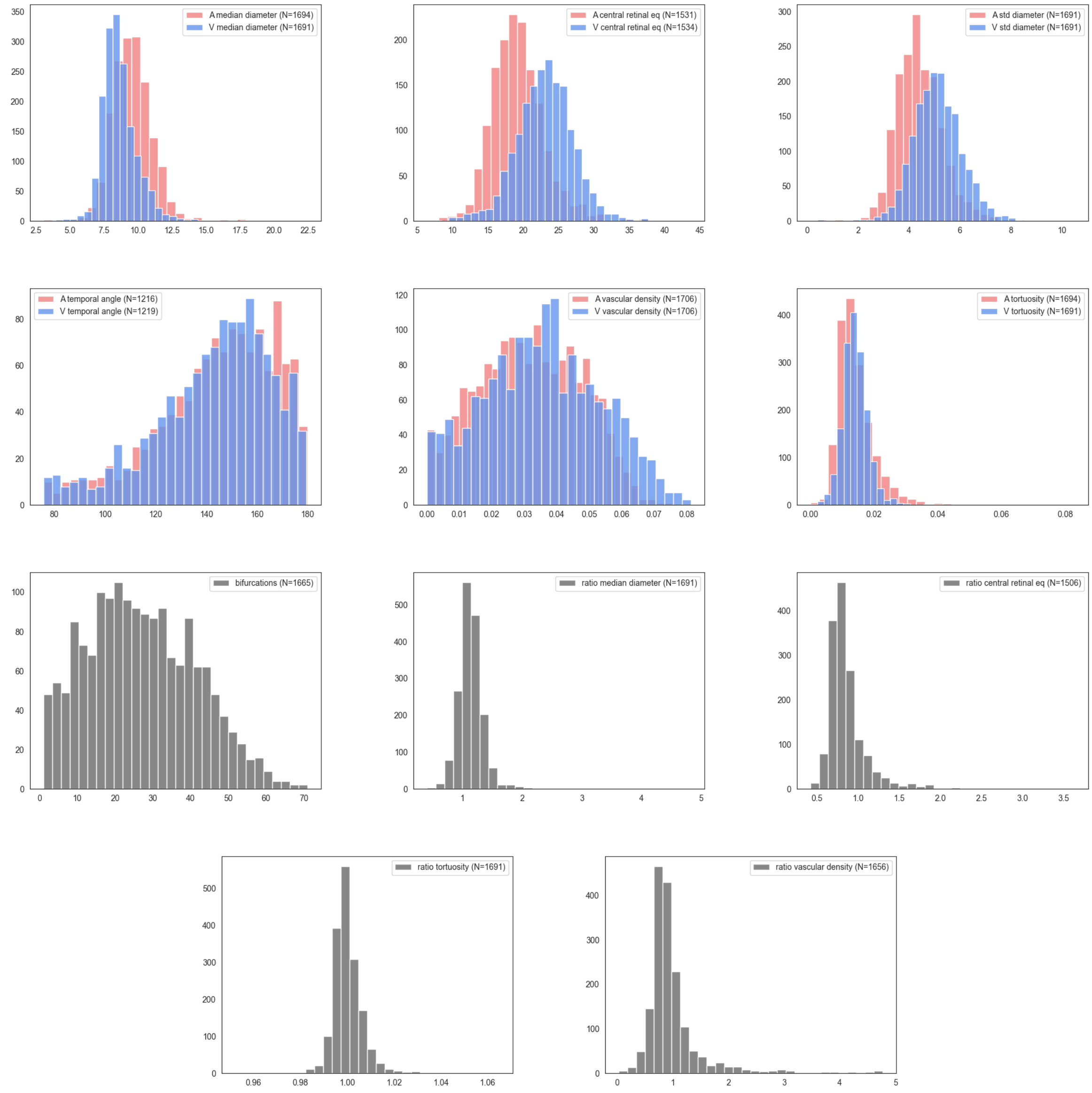


Figure 24: OphtalmoLaus raw distributions of the 17 main IDPs per subject. Outlier values were removed. This outlier removed was especially important for the 'ratio vascular density'.

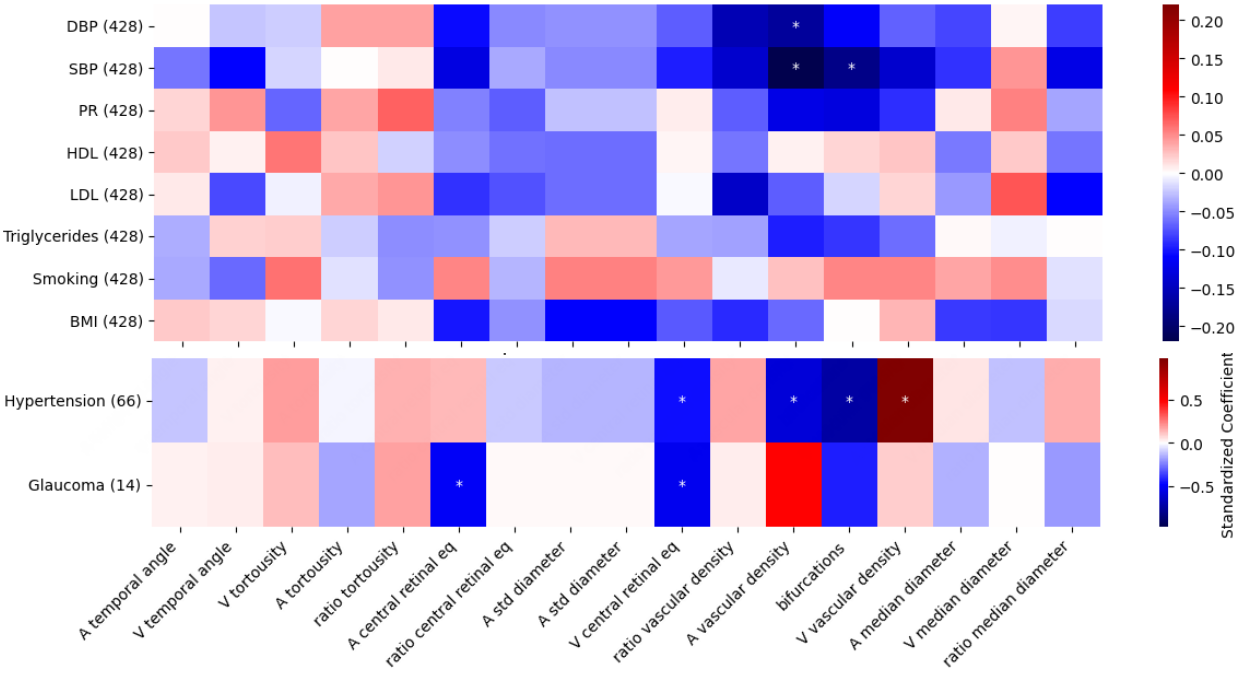


Figure 25: Phenotypic association of IDPs with diseases in the OphtalmoLaus. The x-axis shows IDPs and the y-axis shows diseases. For continuous variables, the numbers in parentheses correspond to the number of subjects with available data, and for binary disease states to the number of cases. Linear (a) and logistic (b) regressions were used for continuous and binary disease states respectively. In all models, phenotypes were corrected for age, sex, eye geometry, and imaging device. The color indicates standardized effect sizes (Standardized Beta) for linear and logistic regressions. Asterisks indicate the level of statistical significance ($* : p < 0.05/Ntests$, $** : p < 0.001/Ntests$, where $Ntests = NIDPs \times Ndiseasetraits$, and $Ndiseasetraits$ is the number of diseases (traits) considered in each panel).

Figure 26 shows the phenotypic correlations between retinal vascular IDPs in the replication and discovery cohorts. From this plot, we can see that the overall phenotypic correlation among the IDPs remains in the replication cohorts, especially for the OphtalmoLaus (Figure 26a).

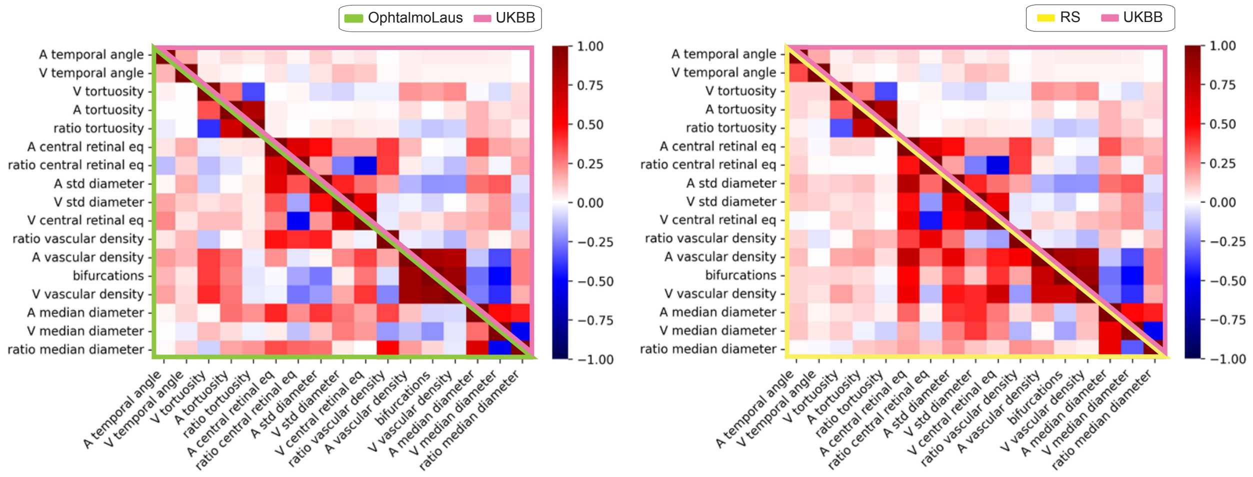


(a) (b)

Figure 26: **a)** Phenotypic correlations between retinal vascular IDPs in the OphtalmoLaus (lower-left green triangle) and UKBB (upper-right pink triangle). **b)** Phenotypic correlations between retinal vascular IDPs in the RS (lower-left yellow triangle) and UKBB (upper-right pink triangle).

#### 17.5 IDPs genetic replication

The GWAS, heritabilities, and gene replication were only applied to the RS, due to the sample size. This includes: a) Manhattan and QQ plots (figures 27 and 28, respectively). b) Benjamini-Hochberg procedure on discovery lead SNPs (figure 29) and genes (figure 31). c) Correlation of effect sizes at the SNP level (figure 30). d) SNP heritabilities values (table 5). e) A table with the significant genes in the RS using the Bonferroni threshold (table 6). For more detailed data refer to the Suppl. Data.

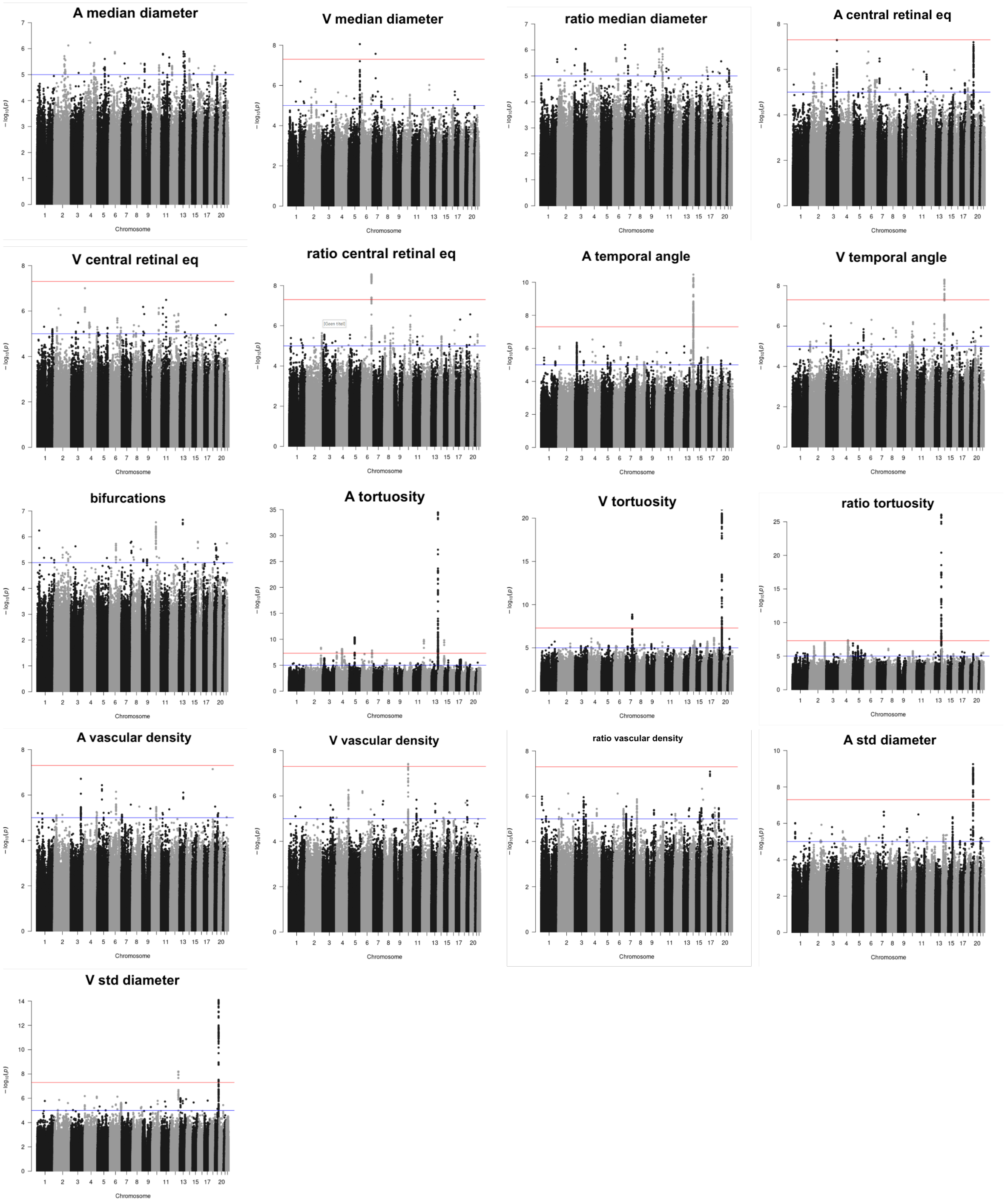


Figure 27: Manhattan plots of the 17 main IDPs used in the main paper from the RS. Each subfigure represents a different IDP. The x-axis displays the positions across the 22 Chromosomes, while the y-axis represents the -log10(p) associated with each SNP. The red line indicates the Bonferroni threshold, above which the dots are considered significantly associated. The blue line indicates the FDR.

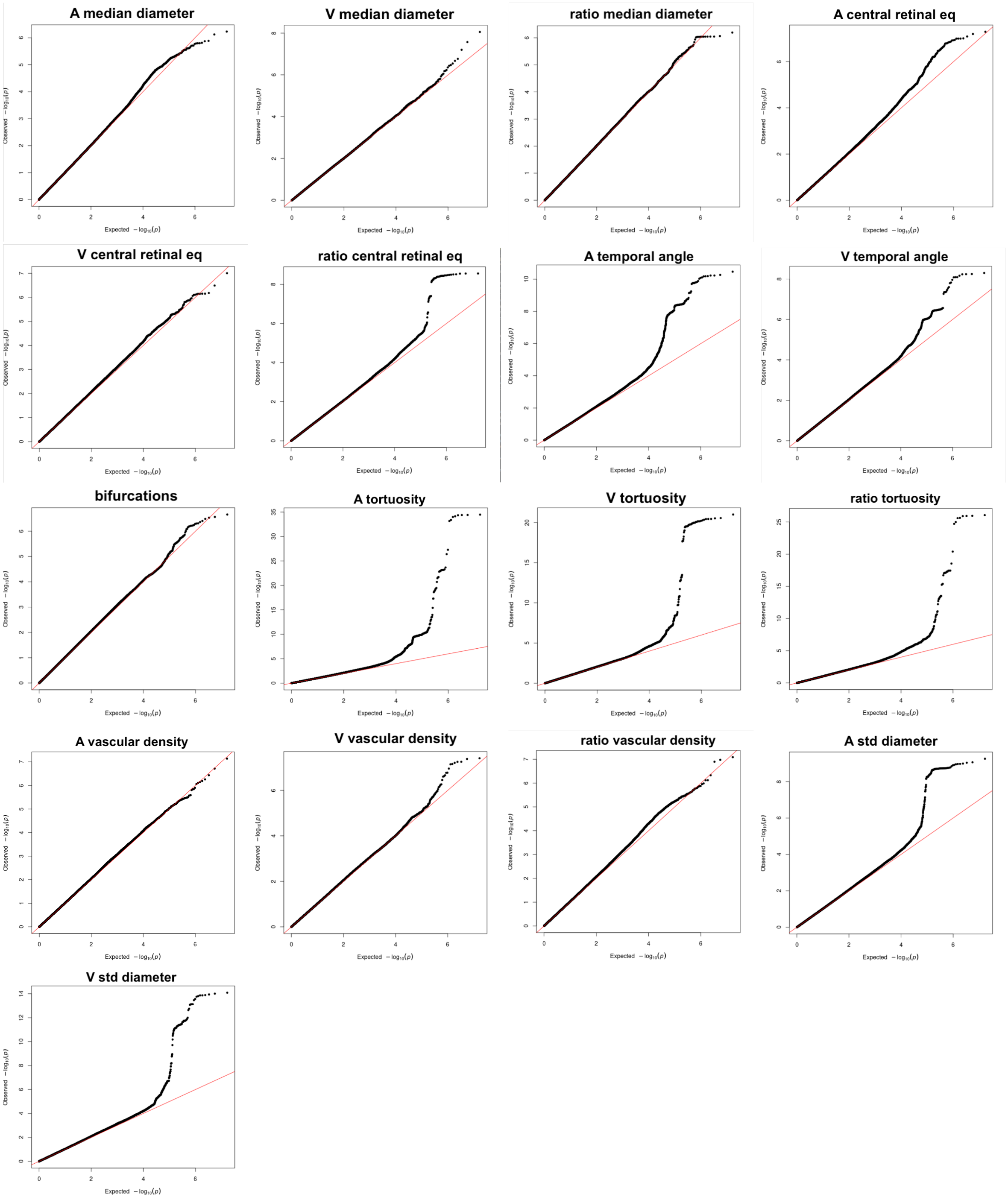


Figure 28: QQ plots of the 17 main IDPs used in the main paper from the RS. Each subfigure represents a different IDP. The x-axis represents expected quantiles under a uniform distribution, while the y-axis represents observed quantiles. Each point on the plot represents a SNP. The alignment between observed and expected quantiles indicates the similarity between the IDPs' distribution and the theoretical uniform distribution.

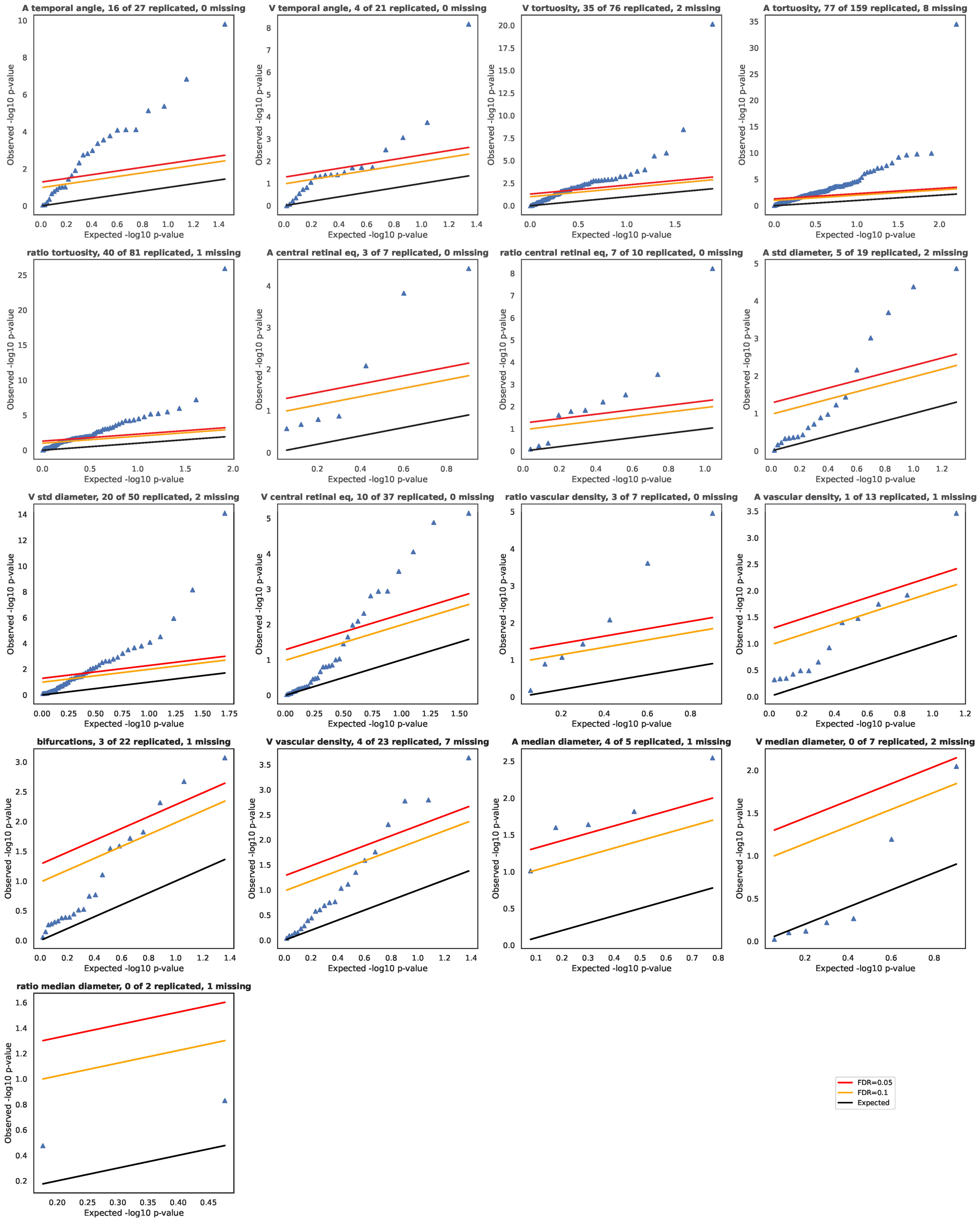


Figure 29: Benjamini-Hochberg procedure on discovery lead SNPs from the UKBB yielded 232 out of 566 discovered hits in the replication cohort, RS, using FDR = 0.05. The label "missing" indicates that these rs were not available in the replication cohort.

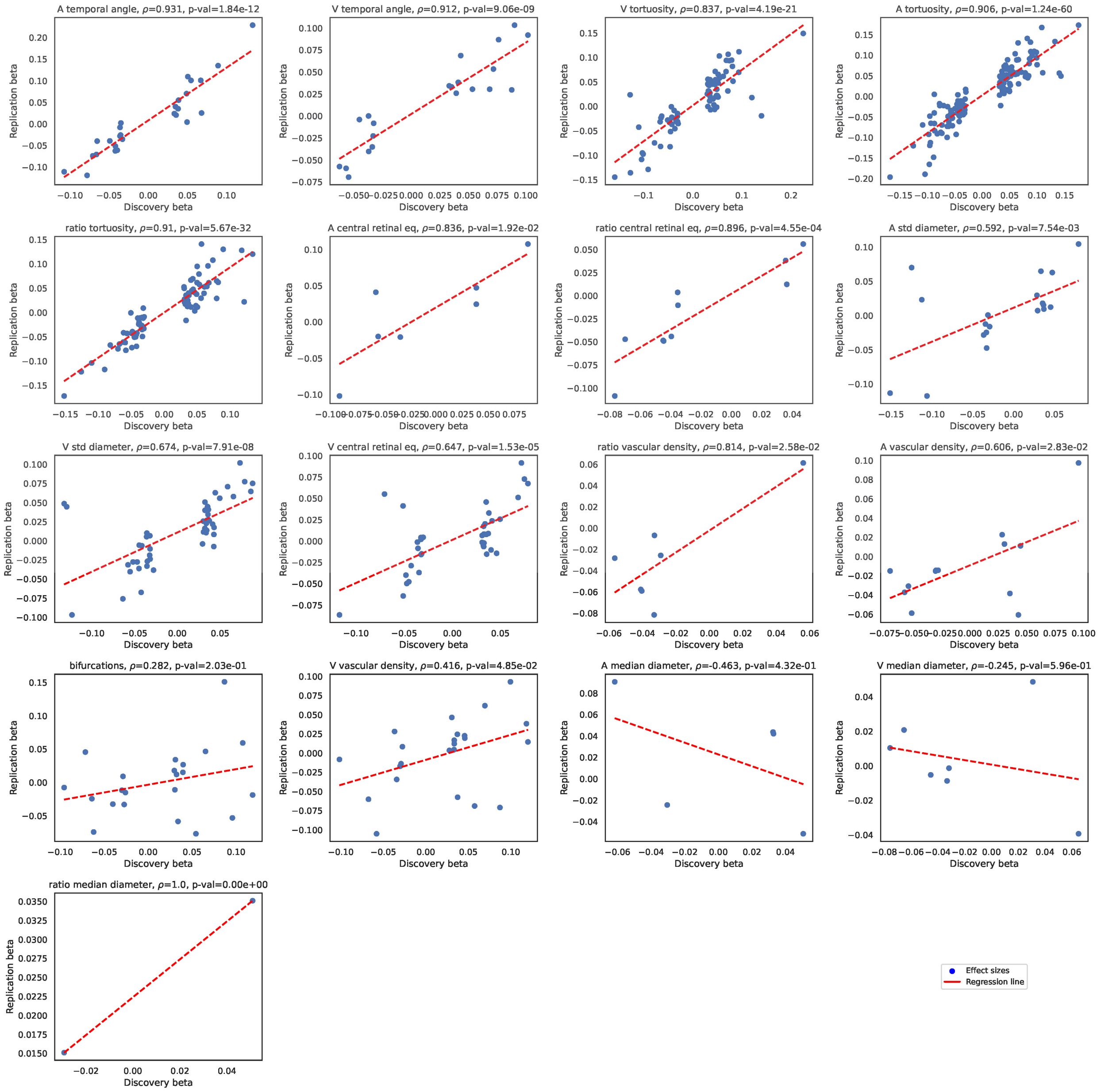


Figure 30: Correlation of effect sizes in the discovery (UKBB) and replication cohort (RS) for the 17 main IDPs, at the SNP level.

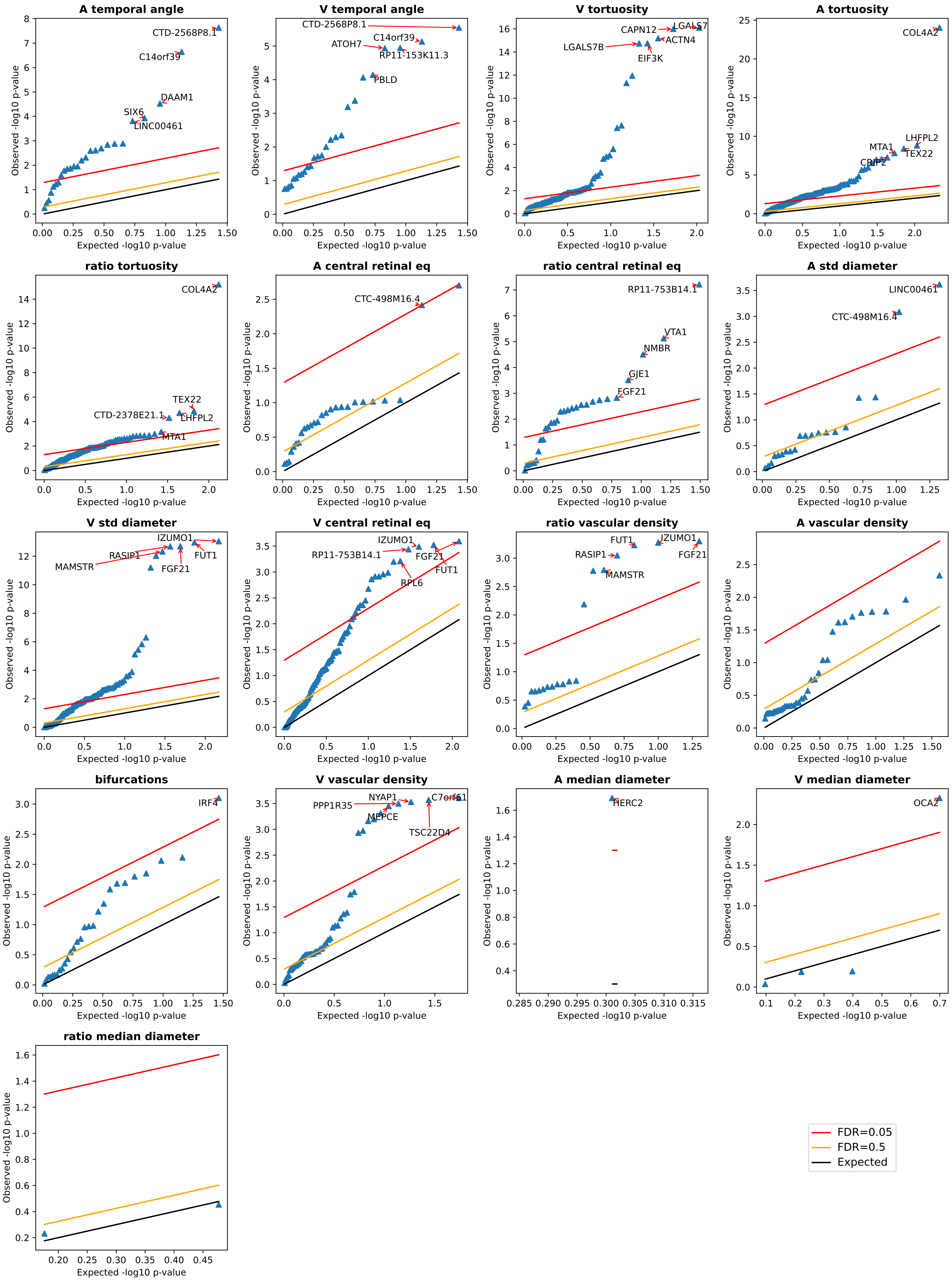


Figure 31: Benjamini-Hochberg procedure on discovery lead genes from the UKBB yielded multiple discovered genes in the replication cohort, RS, using FDR = 0.05. The label "missing" indicates that the corresponding rs of the genes were not available in the replication cohort.

| IDP | $h^2$ | $h^2_{\text{se}}$ | $\lambda_{\text{gc}}$ | Intercept |
|---|---|---|---|---|
| A temporal angle | 0.1455 | 0.0642 | 1.0557 | 1.0344 |
| V temporal angle | 0.0498 | 0.0596 | 1.0345 | 1.0256 |
| V tortuosity | 0.1622 | 0.0573 | 1.0436 | 1.0204 |
| A tortuosity | 0.2235 | 0.0702 | 1.0649 | 1.0454 |
| ratio tortuosity | 0.2022 | 0.0624 | 1.0345 | 1.0232 |
| A central retinal eq | 0.0816 | 0.0593 | 1.0285 | 1.0209 |
| ratio central retinal eq | 0.0781 | 0.0656 | 1.0285 | 1.0176 |
| A std diameter | 0.0802 | 0.0613 | 1.0315 | 1.0219 |
| V std diameter | 0.1782 | 0.0587 | 1.0466 | 1.0206 |
| V central retinal eq | 0.0245 | 0.0544 | 1.0315 | 1.0269 |
| ratio vascular density | 0.0997 | 0.0739 | 1.0315 | 1.0196 |
| A vascular density | 0.0486 | 0.0581 | 1.0225 | 1.0232 |
| bifurcations | 0.0441 | 0.0588 | 1.0195 | 1.0174 |
| V vascular density | 0.0810 | 0.0558 | 1.0285 | 1.0165 |
| A median diameter | -0.0158 | 0.0619 | 1.0016 | 1.0080 |
| V median diameter | 0.1184 | 0.054 | 1.0195 | 1.0025 |
| ratio median diameter | -0.0257 | 0.0614 | 0.9986 | 1.0134 |

Table 5: Main IDPs SNP-based heritability values from RS using LDSR: $h^2$ is the portion of phenotypic variance cumulatively explained by the SNPs. $h^2_{\text{se}}$ indicates the standard error. $\lambda_{\text{gc}}$ is the measure of genomic inflation, it measures the effect of confounding from polygenicity and population stratification acting on the trait. Intercept is the LD Score regression intercept, values close to 1 indicate little influence of confounders, mostly of population stratification.

| **IDP** | **N genes** | **Name genes** |
|---|---|---|
| A temporal angle | 2 | ‘CTD-2568P8.1’, ‘C14orf39’ |
| ratio central retinal eq | 1 | ‘RP11-753B14.1’ |
| A tortuosity | 10 | ‘COL4A2’, ‘LHFPL2’, ‘TEX22’, ‘MTA1’, ‘CRIP2’, ‘CRIP1’, ‘C14orf80’, ‘TMEM121’, ‘TNS1’, ‘CTD-2378E21.1’ |
| ratio tortuosity | 9 | ‘COL4A2’, ‘AC114765.2’, ‘REM2’, ‘AC114765.1’, ‘RBM23’, ‘LRP10’, ‘PRMT5’, ‘HAUS4’, ‘RP11-298I3.5’ |
| V tortuosity | 9 | ‘LGALS7’, ‘CAPN12’, ‘ACTN4’, ‘EIF3K’, ‘LGALS7B’, ‘MAP4K1’, ‘LGALS4’, ‘ECH1’, ‘AC104534.3’ |
| A std diameter | 4 | ‘CTD-2085J24.4’, ‘SLC7A9’, ‘CEP89’, ‘TDRD12’ |
| V std diameter | 9 | ‘IZUMO1’, ‘FUT1’, ‘FGF21’, ‘RASIP1’, ‘MAMSTR’, ‘FUT2’, ‘NTN5’, ‘CA11’, ‘DBP’ |

Table 6: RS significant genes, using Bonferroni threshold. Genes were computed using PascalX.

### 18 Quality control threshold effect on results

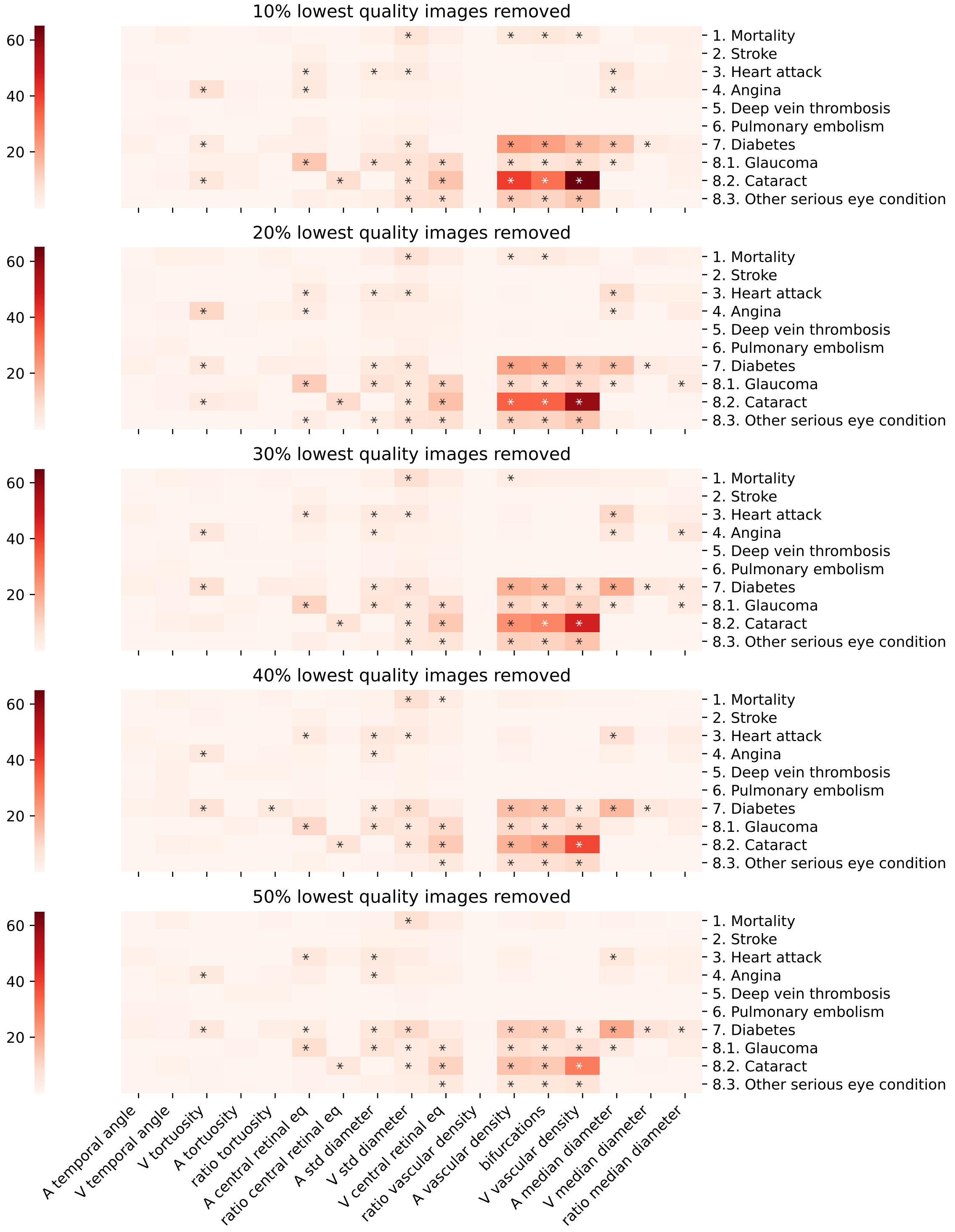


Figure 32: Disease associations in the function of QC stringency. Cox proportional hazard models were used, and Benjamini-Hochberg's false-discovery rate was set to 0.05.

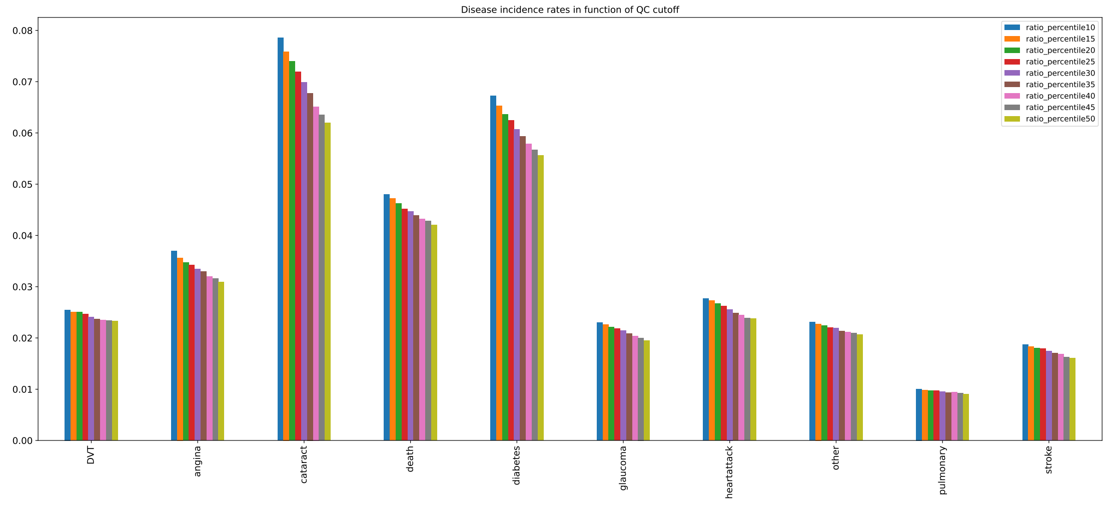


Figure 33: Disease incidence rates (in percent) are plotted in function of QC stringency. We observed a significant decrease in incidence rates, especially cataracts as we removed increasing numbers of bad images.

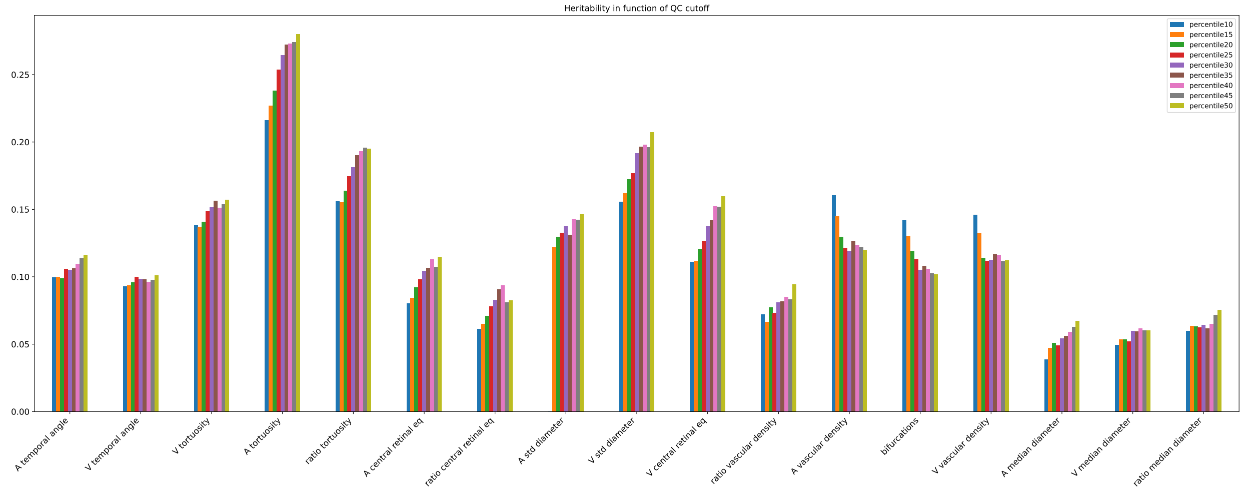


Figure 34: SNP heritabilities as a function of QC. All standard errors were smaller than 0.03, with mean=0.013

### 19 Diseases list UKBB

Medical information regarding the UKBB subjects included in our analysis was available. To explore phenotypically potential risk factors and associations with ocular, vascular, and other diseases, we selected specific information related to these subjects based on previous studies. The list of these diseases used can be found in table 7, with their corresponding identifier "Datafield". Preprocessing was required for certain diseases in the dataset. Specifically, the datafield '2966' representing hypertension, represents the "Age of high blood pressure diagnosis." There are four possible outcomes for this field: "-3" if the individual prefers not to answer, "-1" if they do not know, the actual age of diagnosis if they have hypertension, or an empty value if they don not have hypertension. To binarize this disease, we categorized individuals as hypertensive if they had an age of diagnosis, and as controls if they didn not. Regarding eye diseases, there are four options: having the disease in the right eye, in the left eye, in both eyes, or not having the disease in either eye. To binarize them, we consider individuals as cases if they have the disease in at least one eye, and as controls if they do not have the disease in either eye.

Additionally, for the genetic association analysis with diseases, we utilized the GWAS summary statistics provided by the Neale lab http://www.nealelab.is/uk-biobank. These summary statistics had already been computed for the selected parameters. Researchers interested in further exploring the datafields and files used for both phenotypic and genetic association analyses can refer to "Data Disease information" for additional information.

In summary, our study incorporated medical information from the subjects, additionally, we employed GWAS summary statistics from the Neale lab to investigate the genetic associations with various diseases.

| Category | Disease name | Analysis | Datafield |
|---|---|---|---|
| risk factor | DBP | Linear regression | 4079 |
| risk factor | SBP | Linear regression | 4080 |
| risk factor | PR | Linear regression | 102 |
| risk factor | PWASI | Linear regression | 21021 |
| risk factor | HDL cholesterol | Linear regression | 30760 |
| risk factor | LDL direct | Linear regression | 30780 |
| risk factor | Triglycerides | Linear regression | 30870 |
| risk factor | HbA1c | Linear regression | 30750 |
| risk factor | Alcohol | Linear regression | 1558 |
| risk factor | Smoking | Linear regression | 20161 |
| risk factor | BMI | Linear regression | 21001 |
| vascular | Hypertension | Logistic regression | 2966 |
| eyesight | Amblyopia | Logistic regression | 5408 |
| eyesight | Presbyopia | Logistic regression | 5610 |
| eyesight | Hypermetropia | Logistic regression | 5832 |
| eyesight | Myopia | Logistic regression | 5843 |
| eyesight | Astigmatism | Logistic regression | 5855 |
| ocular | Diabetes-eye | Logistic regression | 5890 |
| ocular | Cataract | Cox model | 4700 |
| ocular | Glaucoma | Cox model | 4689 |
| ocular | Other ED | Cox model | 5945 |
| metabolic | Diabetes | Cox model | 2976 |
| vascular | Angina | Cox model | 3627 |
| vascular | Heart attack | Cox model | 3894 |
| vascular | DVT | Cox model | 4012 |
| vascular | Stroke | Cox model | 4056 |
| vascular | PE | Cox model | 4022 |
| vascular | Atherosclerosis | Cox model | 131380 |
| dementia | AD | Logistic regression | 42020 |
| death | Mortality | Cox model | 40007 |
| vascular | Hypertension | Genetic analysis | 6150_4 |
| risk factor | Smoking status: Never, Current | Genetic analysis | 20116_0, _2 |
| metabolic | Diabetes | Genetic analysis | 2443 |
| vascular | Angina | Genetic analysis | 6150_2 |
| vascular | Heart attack | Genetic analysis | 6150_1 |
| ocular | Cataract | Genetic analysis | 6148_4 |

Table 7: Diseases information used for the IDPs diseases association. Type of diseases with their corresponding UKBB identification, datafield, and the model used.

### References


[1] Mattia Tomasoni, Michael Johannes Beyeler, Sofia Ortin Vela, Ninon Mounier, Eleonora Porcu, Tanguy Corre, Daniel Krefl, Alexander Luke Button, Hana Abouzeid, Konstantinidis Lazaros, et al. Genome-wide association studies of retinal vessel tortuosity identify numerous novel loci revealing genes and pathways associated with ocular and cardiometabolic diseases. *Ophthalmology Science*, page 100288, 2023.

[2] Artem Sevastopolsky. Optic disc and cup segmentation methods for glaucoma detection with modification of u-net convolutional neural network. *Pattern Recognition and Image Analysis*, 27(3):618–624, 2017.

[3] Peter Bankhead, C Norman Scholfield, J Graham McGeown, and Tim M Curtis. Fast retinal vessel detection and measurement using wavelets and edge location refinement. *PloS one*, 7(3):e32435, 2012.

[4] Michael D Knudtson, Kristine E Lee, Larry D Hubbard, Tien Yin Wong, Ronald Klein, and Barbara EK Klein. Revised formulas for summarizing retinal vessel diameters. *Current eye research*, 27(3):143–149, 2003.

[5] Faraz Oloumi, Rangaraj M Rangayyan, and Anna L Ells. A graphical user interface for measurement of temporal arcade angles in fundus images of the retina. In *2012 25th IEEE Canadian Conference on Electrical and Computer Engineering (CCECE)*, pages 1–4. IEEE, 2012.

[6] Harry Pratt, Bryan M Williams, Jae Yee Ku, Charles Vas, Emma McCann, Baidaa Al-Bander, Yitian Zhao, Frans Coenen, and Yalin Zheng. Automatic detection and distinction of retinal vessel bifurcations and crossings in colour fundus photography. *Journal of Imaging*, 4(1):4, 2017.

[7] Harry Pratt, Bryan M Williams, Jae Yee Ku, Charles Vas, Emma McCann, Baidaa Al-Bander, Yitian Zhao, Frans Coenen, and Yalin Zheng. Automatic detection and distinction of retinal vessel bifurcations and crossings in colour fundus photography. *Journal of Imaging*, 4(1):4, 2018.

[8] Álvaro S Hervella, José Rouco, Jorge Novo, Manuel G Penedo, and Marcos Ortega. Deep multi-instance heatmap regression for the detection of retinal vessel crossings and bifurcations in eye fundus images. *Computer Methods and Programs in Biomedicine*, 186:105201, 2020.

[9] Adrian Galdran, André Anjos, José Dolz, Hadi Chakor, Hervé Lombaert, and Ismail Ben Ayed. State-of-the-art retinal vessel segmentation with minimalistic models. *Scientific Reports*, 12(1):1–13, 2022.

[10] Karen Wong, Jeffrey Ng, Anna Ells, Alistair R Fielder, and Clare M Wilson. The temporal and nasal retinal arteriolar and venular angles in preterm infants. *British journal of ophthalmology*, 95(12):1723–1727, 2011.

[11] Mirna Kirin, Reka Nagy, Thomas J MacGillivray, Ozren Polašek, Caroline Hayward, Igor Rudan, Harry Campbell, Sarah Wild, Alan F Wright, James F Wilson, et al. Determinants of retinal microvascular features and their relationships in two european populations. *Journal of hypertension*, 35(8):1646, 2017.

[12] Thomas J MacGillivray, James R Cameron, Qiuli Zhang, Ahmed El-Medany, Carl Mulholland, Ziyan Sheng, Bal Dhillon, Fergus N Doubal, Paul J Foster, Emmanuel Trucco, et al. Suitability of uk biobank retinal images for automatic analysis of morphometric properties of the vasculature. *PLoS One*, 10(5):e0127914, 2015.

[13] Gerald Liew, Jie Jin Wang, Paul Mitchell, and Tien Y Wong. Retinal vascular imaging: a new tool in microvascular disease research. *Circulation: Cardiovascular Imaging*, 1(2):156–161, 2008.

[14] Josef Flammer, Katarzyna Konieczka, Rosa M Bruno, Agostino Virdis, Andreas J Flammer, and Stefano Taddei. The eye and the heart. *European heart journal*, 34(17):1270–1278, 2013.

[15] Tien Yin Wong, Ronald Klein, A Richey Sharrett, Bruce B Duncan, David J Couper, James M Tielsch, Barbara EK Klein, and Larry D Hubbard. Retinal arteriolar narrowing and risk of coronary heart disease in men and women: the atherosclerosis risk in communities study. *Jama*, 287(9):1153–1159, 2002.

[16] Amy McGowan, Giuliana Silvestri, Evelyn Moore, Vittorio Silvestri, Christopher C Patterson, Alexander P Maxwell, and Gareth J McKay. Evaluation of the retinal vasculature in hypertension and chronic kidney disease in an elderly population of irish nuns. *PLoS One*, 10(9):e0136434, 2015.

[17] Xueling Sim, Richard A Jensen, M Kamran Ikram, Mary Frances Cotch, Xiaohui Li, Stuart MacGregor, Jing Xie, Albert Vernon Smith, Eric Boerwinkle, Paul Mitchell, et al. Genetic loci for retinal arteriolar microcirculation. *PloS one*, 8(6):e65804, 2013.

[18] Richard A Jensen, Xueling Sim, Albert Vernon Smith, Xiaohui Li, Jóhanna Jakobsdóttir, Ching-Yu Cheng, Jennifer A Brody, Mary Frances Cotch, Barbara Mcknight, Ronald Klein, et al. Novel genetic loci associated with retinal microvascular diameter. *Circulation: Cardiovascular Genetics*, 9(1):45–54, 2016.

[19] M Kamran Ikram, Sim Xueling, Richard A Jensen, Mary Frances Cotch, Alex W Hewitt, M Arfan Ikram, Jie Jin Wang, Ronald Klein, Barbara EK Klein, Monique MB Breteler, et al. Four novel loci (19q13, 6q24, 12q24, and 5q14) influence the microcirculation in vivo. *PLoS genetics*, 6(10):e1001184, 2010.

[20] Lukas Streese, Lukas Y Brawand, Konstantin Gugleta, Peter M Maloca, Walthard Vilser, and Henner Hanssen. New frontiers in noninvasive analysis of retinal wall-to-lumen ratio by retinal vessel wall analysis. *Translational Vision Science & Technology*, 9(6):7–7, 2020.

[21] Alauddin Bhuiyan, Baikunth Nath, and Kotagiri Ramamohanarao. Detection and classification of bifurcation and branch points on retinal vascular network. In *2012 International Conference on Digital Image Computing Techniques and Applications (DICTA)*, pages 1–8. IEEE, 2012.

[22] WT Longstreth Jr, Emily K Marino Larsen, Ronald Klein, Tien Yin Wong, A Richey Sharrett, David Lefkowitz, and Teri A Manolio. Associations between findings on cranial magnetic resonance imaging and retinal photography in the elderly: the cardiovascular health study. *American journal of epidemiology*, 165(1):78–84, 2007.

[23] Carol Yim-lui Cheung, Yingfeng Zheng, Wynne Hsu, Mong Li Lee, Qiangfeng Peter Lau, Paul Mitchell, Jie Jin Wang, Ronald Klein, and Tien Yin Wong. Retinal vascular tortuosity, blood pressure, and cardiovascular risk factors. *Ophthalmology*, 118(5):812–818, 2011.

[24] Seyedeh Maryam Zekavat, Vineet K Raghu, Mark Trinder, Yixuan Ye, Satoshi Koyama, Michael C Honigberg, Zhi Yu, Akhil Pampana, Sarah Urbut, Sara Haidermota, Declan P O'Regan, Hongyu Zhao, Patrick T Ellinor, Ayellet V Segrè, Tobias Elze, Janey L Wiggs, James Martone, Ron A Adelman, Nazlee Zebardast, Lucian Del Priore, Jay C Wang, and Pradeep Natarajan. Deep learning of the retina enables phenome- and genome-wide analyses of the microvasculature. *Circulation*, 11 2021.

[25] Xi Wang, Qianhua Zhao, Rui Tao, Huimeng Lu, Zhenxu Xiao, Li Zheng, Ding Ding, Saineng Ding, Yichen Ma, Zhaozeng Lu, et al. Decreased retinal vascular density in alzheimer's disease (ad) and mild cognitive impairment (mci): an optical coherence tomography angiography (octa) study. *Frontiers in Aging Neuroscience*, 12:572484, 2021.

[26] Jiong Zhang, Behdad Dashtbozorg, Erik Bekkers, Josien PW Pluim, Remco Duits, and Bart M ter Haar Romeny. Robust retinal vessel segmentation via locally adaptive derivative frames in orientation scores. *IEEE transactions on medical imaging*, 35(12):2631–2644, 2016.

[27] M Kamran Ikram, Frank Jan De Jong, Ewoud J Van Dijk, Niels D Prins, Albert Hofman, Monique MB Breteler, and Paulus TVM De Jong. Retinal vessel diameters and cerebral small vessel disease: the rotterdam scan study. *Brain*, 129(1):182–188, 2006.

[28] Tien Yin Wong, Ronald Klein, Barbara EK Klein, Stacy M Meuer, and Larry D Hubbard. Retinal vessel diameters and their associations with age and blood pressure. *Investigative ophthalmology & visual science*, 44(11):4644–4650, 2003.

[29] Kailimujiang Ahemaitijiangc, Qi Zhange, Dong N Chenf, Chun Zhangb, Fan Lia, Jicong Zhangc, Jost B Jonasa, and Ya X Wanga. Retinal vessel caliber and tortuosity and prediction of 5-year incidence of hypertension. *Journal of Hypertension*, 41:830–837, 2023.

[30] David Rosenbaum, Nadjia Kachenoura, Edouard Koch, Michel Paques, Philippe Cluzel, Alban Redheuil, and Xavier Girerd. Relationships between retinal arteriole anatomy and aortic geometry and function and peripheral resistance in hypertensives. *Hypertension Research*, 39(7):536–542, 2016.

[31] Ryo Kawasaki, Ning Cheung, Jie Jin Wang, Ronald Klein, Barbara EK Klein, Mary Frances Cotch, A Richey Sharrett, Steven Shea, FM Amirul Islam, and Tien Y Wong. Retinal vessel diameters and risk of hypertension: the multiethnic study of atherosclerosis. *Journal of hypertension*, 27(12):2386, 2009.

[32] Ilina Murgan, Sonja Beyer, Konstantin E Kotliar, Lutz Weber, Susanne Bechtold-Dalla Pozza, Robert Dalla Pozza, Aharon Wegner, Diana Sitnikova, Konrad Stock, Uwe Heemann, et al. Arterial and retinal vascular changes in hypertensive and prehypertensive adolescents. *American journal of hypertension*, 26(3):400–408, 2013.

[33] Helena M Pakter, Elton Ferlin, Sandra C Fuchs, Marcelo K Maestri, Ruy S Moraes, Gerson Nunes, Leila B Moreira, Miguel Gus, and Flávio D Fuchs. Measuring arteriolar-to-venous ratio in retinal photography of patients with hypertension: development and application of a new semi-automated method. *American journal of hypertension*, 18(3):417–421, 2005.

[34] Sabrina Koechli, Katharina Endes, Denis Infanger, Lukas Zahner, and Henner Hanssen. Obesity, blood pressure, and retinal vessels: a meta-analysis. *Pediatrics*, 141(6), 2018.

[35] M Kamran Ikram, Frank Jan de Jong, Johannes R Vingerling, Jacqueline CM Witteman, Albert Hofman, Monique MB Breteler, and Paulus TVM de Jong. Are retinal arteriolar or venular diameters associated with markers for cardiovascular disorders? the rotterdam study. *Investigative ophthalmology & visual science*, 45(7):2129–2134, 2004.

[36] M Kamran Ikram, Jacqueline CM Witteman, Johannes R Vingerling, Monique MB Breteler, Albert Hofman, and Paulus TVM de Jong. Retinal vessel diameters and risk of hypertension: the rotterdam study. *hypertension*, 47(2):189–194, 2006.

[37] Robyn J Tapp, Christopher G Owen, Sarah A Barman, Roshan A Welikala, Paul J Foster, Peter H Whincup, David P Strachan, Alicja R Rudnicka, UK Biobank Eye, and Vision Consortium. Associations of retinal microvascular diameters and tortuosity with blood pressure and arterial stiffness: United kingdom biobank. *Hypertension*, 74(6):1383–1390, 2019.

[38] Guangzheng Dai, Wei He, Ling Xu, Eric E Pazo, Tiezhu Lin, Shasha Liu, and Chenguang Zhang. Exploring the effect of hypertension on retinal microvasculature using deep learning on east asian population. *PloS one*, 15(3):e0230111, 2020.

[39] MK Ikram, FJ De Jong, MJ Bos, JR Vingerling, Albert Hofman, PJ Koudstaal, PTVM De Jong, and MMB Breteler. Retinal vessel diameters and risk of stroke: the rotterdam study. *Neurology*, 66(9):1339–1343, 2006.

[40] Hui-Qun Wu, Huan Wu, Li-Li Shi, Li-Yuan Yu, Li-Yuan Wang, Ya-Lan Chen, Jin-Song Geng, Jian Shi, Kui Jiang, and Jian-Cheng Dong. The association between retinal vasculature changes and stroke: a literature review and meta-analysis. *International journal of ophthalmology*, 10(1):109, 2017.

[41] Wenjie Li, Miranda T Schram, Tos TJM Berendschot, Carroll AB Webers, Abraham A Kroon, Carla JH van der Kallen, Ronald MA Henry, Nicolaas C Schaper, Fan Huang, Behdad Dashtbozorg, et al. Type 2 diabetes and hba 1c are independently associated with wider retinal arterioles: the maastricht study. *Diabetologia*, 63:1408–1417, 2020.

[42] Toke Bek. Diameter changes of retinal vessels in diabetic retinopathy. *Current diabetes reports*, 17:1–7, 2017.

[43] Chengxuan Qiu, Mary Frances Cotch, Sigurdur Sigurdsson, Melissa Garcia, Ronald Klein, Fridbert Jonasson, Barbara EK Klein, Gudny Eiriksdottir, Tamara B Harris, Mark A van Buchem, et al. Retinal and cerebral microvascular signs and diabetes: the age, gene/environment susceptibility-reykjavik study. *Diabetes*, 57(6):1645–1650, 2008.

[44] Ronald Klein, Barbara EK Klein, Scot E Moss, Tien Y Wong, Larry Hubbard, Karen J Cruickshanks, and Mari Palta. The relation of retinal vessel caliber to the incidence and progressionof diabetic retinopathy: Xix: The wisconsin epidemiologic study of diabetic retinopathy. *Archives of ophthalmology*, 122(1):76–83, 2004.

[45] Oumayma Khayrallah, Ahmed Mahjoub, Anis Mahjoub, Mohamed Ghorbel, Hechmi Mahjoub, Leila Knani, Fathi Krifa, et al. Optical coherence tomography angiography vessel density parameters in primary open-angle glaucoma. *Annals of Medicine and Surgery*, 69:102671, 2021.

[46] Liang Lv, Mu Li, Xuejiao Chang, Mengxia Zhu, Ying Liu, Ping Wang, and Yan Xiang. Macular retinal microvasculature of hyperopia, emmetropia, and myopia in children. *Frontiers in Medicine*, page 1391, 2022.

[47] Abirami Veluchamy, Lucia Ballerini, Veronique Vitart, Katharina E Schraut, Mirna Kirin, Harry Campbell, Peter K Joshi, Devanjali Relan, Sarah Harris, Ellie Brown, et al. Novel genetic locus influencing retinal venular tortuosity is also associated with risk of coronary artery disease. *Arteriosclerosis, thrombosis, and vascular biology*, 39(12):2542–2552, 2019.

[48] Xiaofan Jiang, Pirro G Hysi, Anthony P Khawaja, Omar A Mahroo, Zihe Xu, Christopher J Hammond, Paul J Foster, Roshan A Welikala, Sarah A Barman, Peter H Whincup, et al. Gwas on retinal vasculometry phenotypes. *Plos Genetics*, 19(2):e1010583, 2023.

[49] Ziqian Xie, Tao Zhang, Sangbae Kim, Jiaxiong Lu, Wanheng Zhang, Cheng-Hui Lin, Man-Ru Wu, Alexander Davis, Roomasa Channa, Luca Giancarlo, et al. igwas: image-based genome-wide association of self-supervised deep phenotyping of human medical images. *medRxiv*, pages 2022–05, 2022.

[50] Brendan K Bulik-Sullivan, Po-Ru Loh, Hilary K Finucane, Stephan Ripke, Jian Yang, Nick Patterson, Mark J Daly, Alkes L Price, and Benjamin M Neale. Ld score regression distinguishes confounding from polygenicity in genome-wide association studies. *Nature genetics*, 47(3):291–295, 2015.

[51] Daniel Krefl and Sven Bergmann. Cross-gwas coherence test at the gene and pathway level. *PLoS Computational Biology*, 18(9):e1010517, 2022.

[52] M Arfan Ikram, Guy Brusselle, Mohsen Ghanbari, André Goedegebure, M Kamran Ikram, Maryam Kavousi, Brenda C T Kieboom, Caroline C W Klaver, Robert J de Knegt, Annemarie I Luik, Tamar E C Nijsten, Robin P Peeters, Frank J A van Rooij, Bruno H Stricker, André G Uitterlinden, Meike W Vernooij, and Trudy Voortman. Objectives, design and main findings until 2020 from the rotterdam study. *Eur. J. Epidemiol.*, 35(5):483–517, May 2020.

[53] Christopher C Chang, Carson C Chow, Laurent Cam Tellier, Shashaank Vattikuti, Shaun M Purcell, and James J Lee. Second-generation PLINK: rising to the challenge of larger and richer datasets. *Gigascience*, 4(1):7, February 2015.

[54] Cristen J Willer, Yun Li, and Gonçalo R Abecasis. METAL: fast and efficient meta-analysis of genomewide association scans. *Bioinformatics*, 26(17):2190–2191, September 2010.
